# Nested pool testing strategy for the reliable identification of individuals infected with SARS-CoV-2

**DOI:** 10.1101/2021.01.03.21249170

**Authors:** Inés Armendáriz, Pablo A. Ferrari, Daniel Fraiman, José M. Martínez, Hugo G. Menzella, Silvina Ponce Dawson

## Abstract

The progress of the SARS-CoV-2 pandemic requires the design of cost-effective testing programs at large scale. To this end, pooling multiple samples can provide a solution. Defining a cost-effective strategy requires the establishment of an efficient deconvolution and re-testing procedure that eventually allows the identifcation of the carrier. Based on Dorfman’s algorithm, we developed an adaptive nested strategy for which we have, for a given prevalence, simple analytic expressions of the optimal number of samples in the starting pool, of the number of partitioning steps (stages) in the optimal path, of the pool sizes in each of these stages and of the expected average number of tests needed to identify the infected individuals. In this paper we analyze the strategy in detail focusing on its practical implementation when there are restrictions that prevent the use of the optimum. More specifically, we analyze how to proceed when the infection prevalence is poorly known *a priori* or when the optimal requires starting with pool sizes that are too large for the reliable detection of an infected sample. The sensitivity of the RT-qPCR assay, the gold standard RNA detection method, is a major concern in the case of SARS-CoV-2: it is estimated that half of the infected individuals give false negative results. Recently, droplet digital PCR (ddPCR) was shown to be 10 − 100 times more sensitive than RT-qPCR, making this technology suitable for pool testing. ddPCR has the added value of providing the direct quantification of the RNA content at the end of the test. In the paper we show how this feature can be used for verification purposes. The analyses and strategies presented here should be useful to those considering the adoption of a pooling approach for RNA detection, particularly, for the identification of individuals infected with SARS-CoV-2.

**Author summary:** The progress of the SARS-CoV-2 pandemic requires the design of cost-effective testing programs at large scale. Running tests on pooled samples can provide a solution if the tests sensitivity is high enough. In the case of SARS-CoV-2, the current gold standard test, RT-qPCR, has shown some limitations that only allow the use of pools with relatively few samples. In this regard, Droplet digital PCR (ddPCR) has been shown to be 10 − 100 times more sensitive than RT-qPCR, making it suitable for test pooling. In this paper we describe a nested pool testing method in which the properties that make it optimal are simple analytic functions of the infection prevalence. We discuss how to proceed in practical implementations of the strategy, particularly when there are constraints that prevent the use of the optimal. We also show how its nested nature can be combined with the direct RNA quantification that the ddPCR test provides to identify the presence of unviable samples in the pools and for self-consistency tests. The studies of this paper should be useful for those considering the adoption of test pooling for RNA detection.

## Introduction

The outbreak caused by the novel Coronavirus, SARS-CoV-2, has spread over almost all countries in the world and running diagnostic tests is required for the analysis of symptomatic individuals. Testing is also a powerful tool used in surveillance at sites of previous or potential outbreaks and for environmental monitoring. The early detection of asymptomatic or pre-symptomatic carriers is crucial to mitigate the spread of the infection, since contact tracing and isolation are required to interrupt the chain of transmission. For this, massive testing campaigns are being implemented, which demand the maximum efficiency in the use of the available resources [1]. Mixing (*pooling*) multiple samples to test them in a single reaction is an option to increase testing capacity reducing costs and time at the same time.

Group or pool testing has been extensively studied for two main applications: (*i*) to identify “defective” units (in the present case, infected individuals) when the prevalence, *p*, defined as the probability of being defective, is relatively low [2]; (*ii*) to estimate *p* [3]. In the present manuscript we mostly focus on the identification problem. This is the key aspect for the expansion of the testing capacity; accepting that a negative result in a single reaction indicates that all the individuals whose samples are included in the tested pool are not infected.

The first proposal of a pooling strategy for this aim was made by Dorfman [4] in 1943. In this strategy, pools are formed containing a number, *m*, of individual samples. One test is run on each pool to detect the presence of a defective or infected sample in it. Assuming that the detection is sensitive enough, the samples in the pools that test negative in this first stage are identified as non-infected. All the samples that belong to pools that test positive are tested individually at a second stage. The identification of the samples that belong to pools that test negative at first is done with only one test. The identification of those that belong to pools that test positive at first requires *m* + 1 tests instead. A large reduction in the number of tests will then be achieved depending on the relative fraction of pools that test positive at the first stage. This scheme is an example of a class of models that are called *adaptative* or *hierarchical* or *sequential*. In this type of models the actions to be taken at any given stage depend on the test results of the previous stages. The two-stage strategy of Dorfman’s was subsequently improved by others [5–7] including more stages to further reduce the number of tests that are necessary to identify the infected samples. In particular, the method of [7], which was recently optimized in [8], achieves an important reduction at the expense of having a random, in principle large, number of stages. Alternatively, *non-adaptive* methods can be applied, in which the way in which the samples are pooled and tested is established before having the result of any test. This definition of the pools *a priori* allows an easy parallelization of the strategy [9] and, depending on the method, can also help to reduce the impact of test errors by including all individual samples in more than one pool. One of the most common such strategies is *array testing* in which each sample is included in at least two pools and if both pools test negative then the individual samples are labeled as not infected, otherwise they are tested individually [10–12]. There are also mixed strategies, called *semi-adaptive*, that try to exploit the advantages of the different types of approaches simultaneously [13]. According to [9], the mixed strategy of [13] is the best two stage strategy in terms of reducing the number of tests per individual. In the present paper we analyze in detail the adaptive nested strategy introduced in [14] which is an extension of Dorfman’s algorithm for more than two stages. In particular, we look at the issues that may arise in practical implementations of the strategy to detect samples infected with SARS-CoV-2.

Various pool testing strategies have been analyzed specifically for the case of SARS-CoV-2. In [15] Dorfman’s algorithm is applied including the use of replicates to check for false negatives or positives. In [16], Dorfman’s strategy is compared numerically with adaptive and non-adaptive methods that use, at some stage, binary splitting. The work in [17] evaluates numerically the performance of two-dimensional array pooling in which the individual samples are organized in an *r × c* array. In [18] a *D*-dimensional array pooling strategy is evaluated assuming that there are very few infected samples among the tested ones. This strategy is being used for large scale testing in Rwanda [19]. In [20], both Dorfman’s and array algorithms are evaluated on practical implementations. In [21] a non-adaptive group testing approach is presented which uses a combinatorial pooling strategy that is based on compressed sensing and is designed to maximize the identification of the positive samples. So far, sample pooling strategies for SARS-CoV-2 nucleic acid detection have mostly been explored for the current gold standard test, RT-qPCR [22], finding that, for large enough viral loads, the identification of a single infected individual is possible in pools of up to 100 samples (1 infected + 99 negative samples). Despite the initial excitement, it is accepted that in RT-qPCR the target can go unidentified if present in small amounts, and that samples frequently contain inhibitors of the PCR reaction [23]. Moreover, diagnostic sensitivity in symptomatic patients was reported to be in the range of 60%–90% [24]. Thus, for samples with low viral loads, the pooling associated dilution will further reduce the detection of infected individuals to unacceptable levels. The risk of increasing the number of false negative results has raised concerns and delayed the approval of group testing protocols, limiting pool sizes to single digit numbers. This has reduced the impact of pooling strategies and, consequently, the expansion of the world testing capacity.

Recently, great improvements in the sensitivity and specificity of SARS-CoV-2 RNA detection have been introduced by using droplet digital PCR (ddPCR). In ddPCR the sample is emulsified with oil to split the reaction in thousands of droplets [25]. Each droplet works as a tiny reactor, reducing the chance of interferences and favoring the detection of even a single molecule of the target nucleic acid. ddPCR provides orders of magnitude more precision and sensitivity than RT-qPCR and allows for the direct quantification of the nucleic acid content in the samples. A direct comparison in samples with low RNA concentration and variable amounts of inhibitors, ddPCR was shown to be up to 500 times more sensitive than RT-qPCR [26, 27]. Other technologies like Next Generation Sequencing were reported to be equally-effective as ddPCR at improving the limit of the detection of SARS-CoV-2 nucleic acid [28]. The implementation of new technologies like ddPCR would remove therefore the major barrier for the adoption of group testing strategies. This might in turn dramatically improve the current world testing capacity soon, raising the need to rapidly obtain efficient algorithms for pool testing [29]. Here we develop the adaptive nested strategy introduced in [14] to address the issues that may arise in practical implementations to detect the presence of SARS-CoV-2 RNA.

The strategy introduced in [14] has simple analytic expressions for the optimal number of stages and pool sizes and for the expected number of tests that will allow the identification of the infected individuals in a population as functions of the infection probability, *p*. In the present paper we analyze how to proceed when *p* is not known *a priori* or when there are constraints that prevent the optimal strategy from being used. The analytic nature of the results allows these issues to be handled easily, particularly, to find the optimal strategy when there is an upper bound on the number of samples that can be mixed in a pool. As shown in the paper, the strategy can also be easily implemented in parallel and, thanks to the analytic expressions, the expected number of tests per stage can be estimated *a priori* which allows the experimenter to make an informed decision on the best way to proceed given the limitations of their set-up. The nested nature of the strategy, on the other hand, allows for result verification when using a type of test that gives a direct quantification of the detected RNA content, such as ddPCR. Regarding cost reduction, for small enough *p*, the expected number of tests per individual of our optimal strategy is ∼ 3*p* log(1*/p*)*/* log(3) ≈ 2.7*p* log(1*/p*), which is similar to that of the three dimensional array testing studied in [18] and slightly larger than that of the mixed strategy of [13], ∼ *p* log(1*/p*)*/* log^2^(2) ≈ 2.1*p* log(1*/p*) or of the binary splitting one, ∼ *p* log(1*/p*)*/* log(2) ≈ 1.4*p* log(1*/p*). The cost reduction of the latter, however, is achieved at the expense of having an unknown number of stages,, which is impractical for clinical laboratories due to the pressure imposed by fixed delivery time frames. The order of magnitude of the reduction for small *p* is mainly determined, in all three cases, by the *p* log(1*/p*) factor, which is the same for all. The simplicity of our strategy and the advantages of its nested nature make it a very valuable option to faithfully detect viral RNA saving costs at the same time. We think it can be very useful to minimize the variable costs of SARS-CoV-2-carrier detection and optimize the use of resources in testing facilities. An interface for interested users of the method will be made available shortly at https://wp.df.uba.ar/pooling.

## Methods

### Nested pooling method: description and main formulas

We briefly describe hereby the strategy that we analyze in the present paper quoting the formulas that we will use. More details are provided in S1 Appendix, Sec. 1. The strategy, introduced in [14], consists in iterating Dorfman’s [4] to more than two stages using nested pools. This means that the pools of each stage are obtained as a partition of the pools that tested positive at the previous one. All the pools of a given stage contain the same number of samples. For the definition and optimization of the strategy we work under the assumption that if there is at least one infected sample in a pool the test detects its presence. We also assume that the test does not give “false” detections of infected samples. We discuss in the Results Section how to consider the existence of possible false negatives in pool testing.

The strategy is characterized by the number of stages, (*k* + 1), and the sequence of pool sizes, *m* = (*m*_1_, …, *m*_*k*_), where *m*_1_ *> … > m*_*k*_ *>* 1 and *m*_*j*_ is a multiple of *m*_*j*+1_ for all *j*. At the first stage, pools with *m*_1_ samples are tested. Those that test positive are split into pools of size *m*_2_ and are tested at a second stage. The procedure is repeated until the *k* + 1-th stage. At this last stage all the samples contained in pools that tested positive at the *k*-th stage are individually tested, *i.e*., *m*_*k*+1_ = 1. We illustrate the strategy with *k* = 2 and *m* = (9, 3) in Fig. 1.

**Fig 1.**
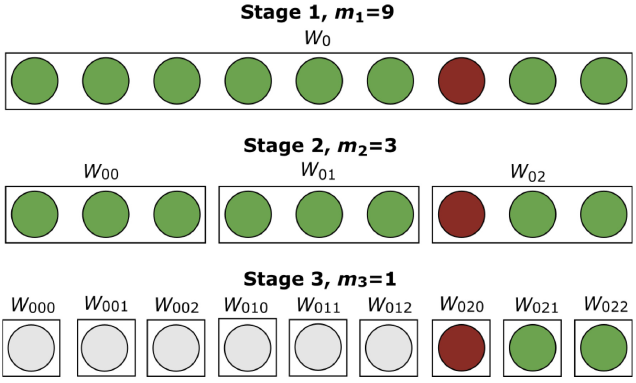
Schematic depiction of the nested pool testing strategy. In the figure the strategy with *k* = 2 and *m* = (9, 3) is illustrated when applied to one pool labeled, *W*_0_. The circles represent individual samples and the rectangles around the colored ones represent pools. The red circles correspond to infected samples. The circles corresponding to the non-infected ones are either green (when they belong to a pool that is being tested at the corresponding stage) or grey (when they have been identified as non-infected at a previous stage). At the first stage all 9 samples belong to the same pool, *W*_0_, and are tested together. Given that there is an infected sample in *W*_0_, the test identifies the pool as infected. Thus, *W*_0_ is divided into 3 sub-pools, *W*_00_, *W*_01_, *W*_02_, of 3 samples each that are tested at the second stage. After these tests, *W*_00_ and *W*_01_, are identified as not infected so that the samples in them are not tested at the third stage. *W*_02_, on the other hand, is identified as infected and all its samples are tested individually at the last stage. As discussed in the main text and in S1 Appendix, Sec. 1, a label, 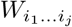, can be assigned *a priori* to the individual samples and the pools of all stages, regardless of whether they are going to be tested as such or not. As illustrated in the figure, it is characterized by a sequence of as many subscripts, *i*_1_, …, *i*_*j*_, as stage number with, *i*_1_, the subscript that identifies the initial pool (in the figure, *i*_1_ = 0).

Given the infection probability, *p*, the optimal strategy is characterized by the values, *k* and *m*, that minimize the cost, *D*_*k*_(*m, p*), defined as the expected number of tests per individual, *ET*_*k*_*/m*_1_. The optimal value of *k* increases with decreasing *p*. Thus, it only makes sense to use our pooling strategy if [14]:

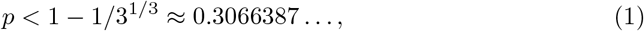

which is the equivalent of the upper bound of Dorfman’s for integer-sized pools. As described in S1 Appendix, Sec. 1, we obtained [14] analytic expressions for the expected number of tests per initial pool at each stage, *j*:

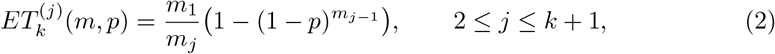

for the expected total number, 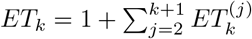, and for the cost of the strategy:

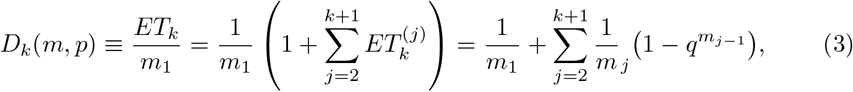

where *q* = 1 − *p* is the probability that an individual is not infected.

Among the universe of nested strategies, those with *m* = (*µ*^*k*^, …, *µ*) and *µ* an integer larger than one are of special interest. In particular, we proved in [14] that, for most values of *p*, the optimal nested strategy is of this form with *µ* = 3 (see S1 Appendix, Sec. 1). For this reason in this paper we mainly focus on strategies with *m* = (3^*k*^, …, 3). Restricting the analysis to strategies with *m* = (*µ*^*k*^, …, *µ*), *µ >* 1, we proved in [14] that the *k* that optimizes the cost, *D*_*k*_(*m, p*), is *k* = *k*_*µ*_(*p*) with:

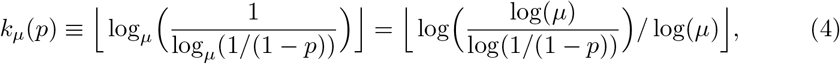

where ⌊ζ⌋ is the largest integer, *n*_*ζ*_, such that *n*_*ζ*_ ≤ *ζ*. We further proved in [14] that setting *k* = *k*_3_(*p*), with *k*_3_(*p*) given by Eq. (4) and *µ* = 3, the cost, 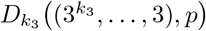 is of the same order of magnitude, *O* (*p* log(1*/p*)), as the cost of the optimal strategy for that *p*. Thus, a strategy with *m* = (3^*k*^, 3^*k*−1^, …, 3) and *k* = *k*_3_(*p*) will provide good results (*i.e*., an expected number of tests per individual of the same order of magnitude as the one of the optimal nested strategy) for any *p <* 1 − 1*/*3^1*/*3^.

We also found in [14] that when *m* = (*µ*^*k*^, …, *µ*) the standard deviation of the number of tests per individual can be written as:

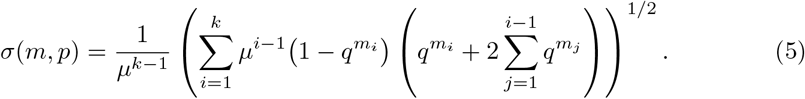

### Nested pooling method: stochastic numerical simulations

In this paper we show the result of stochastic simulations of the strategy. Each stochastic simulation is characterized by the infection probability, *p*, the number of stages before the individual testing, *k*, the sequence of pool sizes, (*m*_1_, *m*_2_, …, *m*_*k*_), and the number of realizations or, equivalently, the number of initial pools, *N*_*w*_. Most of the simulations shown in the paper have *N*_*w*_ = 1000. To run a simulation a sequence of 0s and 1s of length *m*_1_*N*_*w*_ is generated choosing the 1s with the desired infection probability, *p*. To this end a sequence of *m*_1_*N*_*w*_ (pseudo) random numbers is generated with uniform probability over [0, 1). The 1s are then assigned to the positions in the sequence for which the corresponding random number is less or equal *p*. These elements of the sequence correspond to the infected samples. The rest of the positions are set equal to zero and correspond to the non-infected ones. The samples in each initial pool are chosen sequentially along the *m*_1_*N*_*w*_-long sequence. The strategy is applied as previously explained: given a stage, *j*, and a pool with *m*_*j*_ samples, only the pools that contain a 1 in their sequence are tested at stage, *j* + 1, for which they are subdivided into *m*_*j*_*/m*_*j*+1_ pools of *m*_*j*+1_ samples each. As in the case of the first stage, the samples in each subpool that is tested at the *j* + 1 stage are chosen sequentially along the sequence that characterizes the pool of the *j*-th stage from which they derive. This way of organizing the simulation uses the labeling of pools and samples that is illustrated in Fig. 1 and described in detail in S1 Appendix, Sec. 1. In brief, if we order the initial individual samples in a row, as done in Fig. 1, and define *a priori* the pools of the *j*-th stage as the sets containing *m*_*j*_ subsequent samples along this row (the rectangles in Fig. 1), we associate (*k* + 1) subscripts, *i*_1_*i*_2_…*i*_*k*+1_, to each individual sample that serve to uniquely identify the pools each of them belongs to at any given stage. This is apparent in Fig. 1 where the individual samples are represented along a horizontal line with the first one starting from the left being labeled by the subscripts 000 and the last one by 022. In this case we are considering only one initial pool, *W*_0_, and that is why the first subscript of each individual sample is *i*_1_ = 0. If the number of first stage pools (of size *m*_1_) is *N*_*w*_, then 0 ≤ *i*_1_ ≤ *N*_*w*_ − 1 and 0 ≤ *i*_*j*_ ≤ *m*_*j*−1_*/m*_*j*_ for 2 ≤ *j* ≤ *k*. Let us describe for simplicity the case in which *m*_*j*−1_*/m*_*j*_ = 3. Let us consider an individual sample with subscripts, *i*_1_…*i*_*k*+1_. Then, *i*_1_ gives the order along the row (starting from 0) of the first stage pool that the sample belongs to. The order of the second stage pool it belongs to is 3*i*_1_ + *i*_2_, that of the third one is 3^2^*i*_1_ + 3*i*_2_ + *i*_3_, and so on. In particular, the position along the row (starting from 0) of the individual sample with subscripts is *i*_*k*+1_ + 3*i*_*k*_ + … + 3^*k*−1^*i*_2_ + 3^*k*^*i*_1_. This labeling is also illustrated in Fig. 1 of S1 Appendix, Sec. 1.

Some of the histograms obtained with the simulations also include theoretical computations. These are done counting exhaustively the combinations that lead to the various end results in terms of the number of tests, for numbers that do not exceed a certain value (see S1 Appendix, Sec. 1 for more details).

## Results and Discussion

### Properties of the nested strategies with *m* = (3^*k*^, …, 3)

We illustrate in this Section the main properties of the nested strategies described in Methods that have *k* + 1 stages and a sequence of pool sizes, *m* = (3^*k*^, …, 3). To this end we use analytic results and stochastic numerical simulations.

### Analytic results

For each infection probability, *p*, the optimal strategy of the type *m* = (3^*k*^, …, 3) is determined by the number of stages, *k*. For each *p*, the value, *k*, that characterizes such optimal strategy is given by the function, *k*_3_(*p*), (Eq. (4) with *µ* = 3). *k*_3_(*p*) is a staircase function of *p* that increases with decreasing *p* as shown in Fig. 2 (a).

**Fig 2.**
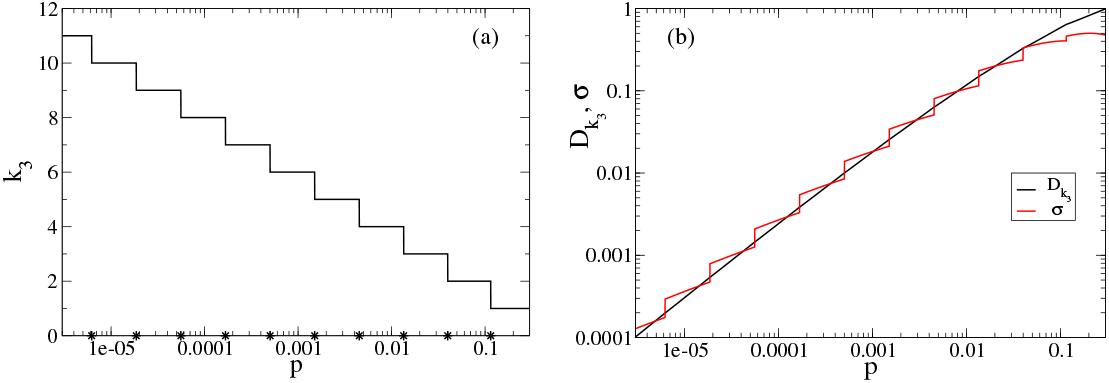
Properties of the nested pool testing strategy. (a) Optimal number of stages, *k*_3_, given by Eq. (4) with *µ* = 3, as a function of ∼ *p. k*_3_(*p*) is a piece-wise constant function with discontinuities at the probabilities indicated with stars on the horizontal axis that increases with decreasing *p*. Thus, the optimal number of stages increases as the infection probability gets smaller. (b) Expected value, 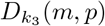 (black curve), and standard deviation, *σ*(*m, p*) (red curve), of the number of tests per individual for strategies of the form 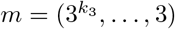 with *k*_3_(*p*) given by Eq. (4). 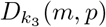 is continuous and *σ* is not, but they both stay close to one another and decrease monotonically with *p*.

We show in Fig. 2 (b) the expected number of tests per individual, *D*_*k*_ (black, Eq. 3), and the standard deviation of this number *σ*(*m, p*) (red, Eq. (5)) for strategies with *m* = (3^*k*^, …, 3) and *k* = *k*_3_(*p*), the optimal number of stages depicted in Fig. 2 (a). The fact that *D*_*k*_ ∼ *σ*(*m, p*) could be worrisome, but is not. If the strategy is applied several times, the average number of tests per individual will approach the expected value, *D*_*k*_ = *ET*_*k*_*/µ*^*k*^. For example, if one performs 100 independent realizations of the strategy with *k* = *k*_*µ*_ given by Eq. (4) for the given *p*, we can expect that the average will be within 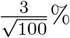 of 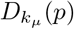 (with probability ≈ .99, by the central limit theorem). This means that the expected value, *D*_*k*_, gives a good estimate of the average number of tests per individual once the strategy has been applied for a while. Furthermore, as discussed in what follows, the relatively large deviation is mainly due to the many instances in which the number of tests is smaller than the expected value.

### Stochastic simulations

We now analyze the interplay between the expected value and the standard deviation of the number of tests per individual with stochastic numerical simulations of the strategy with *m* = (3^3^, 3^2^, 3) for values, *p*, for which it is optimal. The simulations are done as explained in Methods. To this end, we start with 1000 initial pools for each *p, i.e*., we are considering a population of 27,000 individuals within which we choose the infected ones with probability, *p*. Fig. 3 shows the histogram (as fraction of occurrences) of the number of tests performed per pool, using a log scale in the vertical axis, for simulations with *p* = 0.02 (a), *p* = 0.0398 (b) and *p* = 0.0135. The number of infected individuals (the prevalence or fraction of infected individuals) of the simulations were: 544 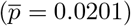 (a); 1073 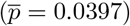 (b) and 369 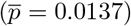 (c). The analytical results derived for some of the cases as explained in S1 Appendix, Sec. 1, are also shown with green bars.

**Fig 3.**
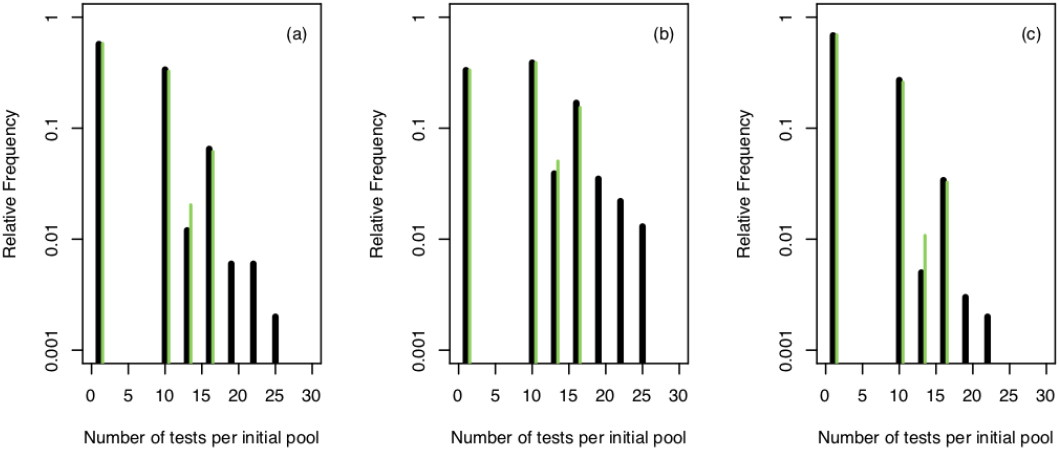
Stochastic simulations of the nested strategy with *k* = 3 and *m* = (3^3^, 3^2^, 3). We show the results obtained when the strategy is applied to 1000 pools with 27 samples each. The figures show histograms (as fraction of occurrences and in log scale) of the number of tests performed per pool. (a) *p* = 0.02 (544 infected samples) (b) *p* = 0.0398 (1073 infected samples) (c) *p* = 0.0135 (369 infected samples). Green bars correspond to the theoretical values that we computed (see S1 Appendix, Sec. 1). The apparent mismatch between theory and simulations is solely due to fluctuations and disappears as the number of initial pools is increased.

We observe in Fig. 3 (a) that approximately 58% (theoretical value (1 − 0.0201)^27^) of the 27-sample pools require only one test, *i.e*., contain no infected samples. It implies that 15,660 individuals are expected to be reported as negative using only 580 tests. This is the main reason for the large reduction in the average number of tests per individual (∼ 0.2, instead of 1) that need to be performed in this example. This becomes apparent if we contrast the histogram of Fig. 3 (a) with the average number of tests per pool, ∼ 5.4. The second most frequent situation in Fig. 3 (a) (with ∼ 34% of the instances) is that in which the number of tests performed per pool is 10. This corresponds to initial pools with only one infected sample or to the few ones with more than one infected sample that remain in the same pool until the *k*-th stage (see S1 Appendix, Sec. 1). In such a case, at any given stage, only one pool “makes it” to the following stage. Thus, the total number of tests that have to be performed for each of these pools is:

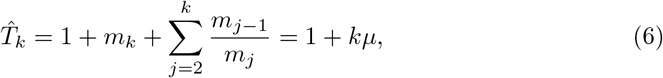

and the number of tests per individual can be written as:

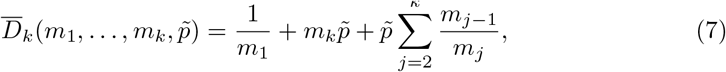

with 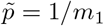. The latter is equal to the expected number of tests per individual for realizations with at most one infected sample per initial pool if we set 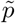 equal to the infection prevalence, 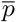. It also coincides with the linearization of Eq. (3) replacing, 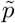, by the infection probability, *p* (see S1 Appendix, Sec. 1). We proved [14] that the cost of the optimal nested strategy is smaller than the one given by Eq. (7). This is reflected in the example of Fig. 3 (a).

The comparison of the three examples of Fig. 3 provides an explanation for the exchange in the relative order of the mean and standard deviation of the number of tests that is apparent in Fig. 2 (b). Let us call, *p*_*µ, ℓ*_, the probability, *p*, such that log_*µ*_(1*/* log_*µ*_(1*/*(1 − *p*)) =, *ℓ* ∈ ℕ, *i.e*., by Eq. (4) *p*_*µ, ℓ*_ is the probability at which the strategy 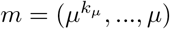 switches from *k*_*µ*_ =, *ℓ* to *k*_*µ*_ =, *ℓ* + 1. In particular, the values, *p*_3,*k*_ with 2 ≤ *k* ≤ 11, are indicated with stars on the horizontal axis of Fig. 2. The infection probabilities in Figs. 3 (b) and (c) satisfy, respectively, *p*, ≲*p*_3,3_ and *p* ≳ *p*_3,4_, and the one in Fig. 3 (a) is in between the two. As may be observed in Fig. 2 (b), while the mean is larger than the deviation for *p*, ≲ *p*_3,3_, the relationship is reversed for *p* ≳ *p*_3,4_. The comparison of Figs. 3 (a)–(c) shows that, as *p* decreases within the range for which *k*_3_ = 3 is optimal, the fraction of initial pools that require the largest number of tests decreases as well while the fraction of those that require only one test (*i.e*., no infected samples), grows. The increasing ratio, *σ/D*_3_, can then be related to an increasing fraction of instances with no infected samples that are resolved with only one test. The reason why *σ* stays relatively “large” as this happens is easy to understand if we assume that the initial pools can only have 0 or 1 infected sample. In such a case, those with no infected samples are solved with a single test while the others require a total number of tests, 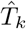, given by Eq. (6). Let us define the stochastic variable, *X*, such that *X* = 0 if the pool contains no infected samples and *X* = 1 otherwise, and let us call, 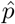, the probability that *X* = 1. The mean and standard deviations of *X* are 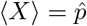 and 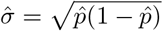 and satisfy 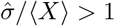 for 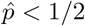 with 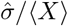 increasing for decreasing 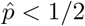. Thus, the ratio increases because the deviation approaches zero more slowly than the mean. This translates into a similar behavior for the number of tests for which the mean and standard deviations are, respectively, 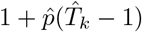 and 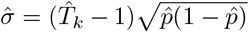. From the point of view of the number of tests it means that the “worst” pools (those with exactly one infected sample) are solved with a number of tests given by Eq. (6) which is clearly smaller than the number of samples in the initial pool, *µ*^*k*^, for *µ* = 3 and *k* ≥ 2.

The optimal strategy of the form (*µ*^*k*^, …, *µ*) for a given *p* is such that *pµ*^*k*^ *<* 1. Thus, initial pools with more than 1 infected sample have a (pool) fraction of infected samples larger than *p*. In such a case, the best case scenario is the one where the infected samples fall after each partition in the same subpool up to the *k*-th stage. Such a situation becomess less likely as the number of infected samples gets larger (*e.g*., 4 or 3, as in the discussed examples). It does occur, however, for 2 infected samples. This is illustrated in Fig. 4 where we show histograms of the number of infected samples per pool for the same stochastic simulation as the one of Fig. 3 (a). There it can be observed that ∼ 6% of the 1545 pools that are “tested” at the third-stage had two infected samples. The pools holding more than one infected sample at the *k*-th stage help are expected to reduce the average number of tests per individual needed to identify the infected ones. They therefore contribute to the inequality between the actual expected value of the number of tests per individual and the one where no more than one infected sample is present in each pool at all stages (see S1 Appendix, Sec. 1).

**Fig 4.**
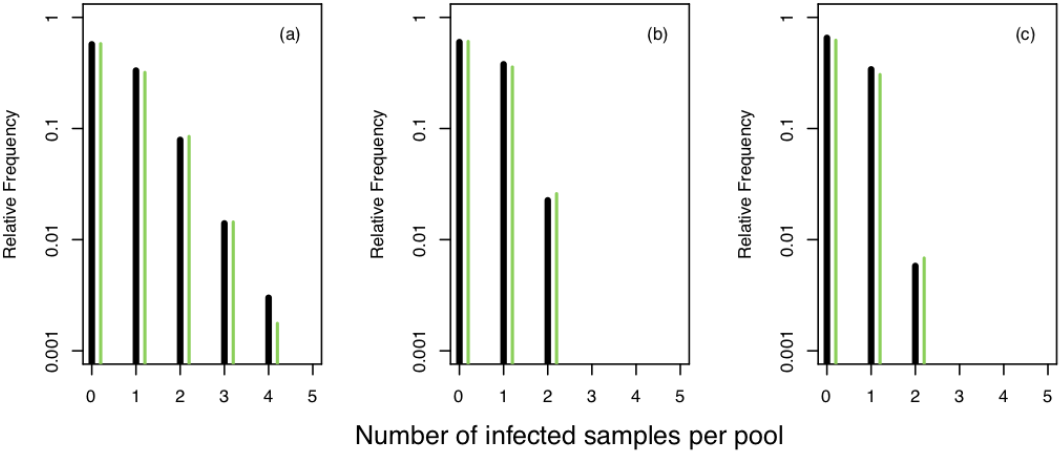
Number of infected samples in a pool. Histogram (as fraction of occurrences in log scale) of the number of infected samples per pool at the first (a), second (b) and third (c) stages for the same example as in Fig. 3 (a). Vertical axis in log-scale. Green bars correspond to the theoretical values that we computed (see S1 Appendix, Sec. 1).

*Based on all the analyses described so far, we then propose to use a strategy of the form*. 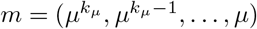 *with µ* = 3 *and k*_*µ*_ *given by Eq. (4) for any value of p, including the smallest ones, whenever possible*.

### Practical implementation of the nested strategy: constraints and runs in parallel

In this Section we present a series of analyses to determine the best nested strategy when the use of the optimal one described so far is prevented by constraints or when the strategy is applied in parallel on a given number of pools at the same time. Namely, two problems that we may encounter in a practical implementation of the nested pool strategy is uncertainty in the estimate of *p* and an incompatibility between the optimal pool sizes and the largest number of samples that can be pooled to obtain reliable results. Even though *p* can be estimated when the strategy is applied allowing and the strategy can be retuned (see S1 Appendix, Sec. 2), in this Section we study the cost reduction that is obtained when using our nested strategy outside its region of optimality. Next we provide a solution for cases in which the maximum number of samples that can be pooled is below the first stage pool size, *m*_1_, of the optimal nested strategy for the given *p*. Finally, the performance of the nested pool strategy is analyzed when the test is run in parallel on several pools at the same time. We compare, in particular, the expected number of pools to be tested at any given stage with those tested at the first one.

### Pooling tests under unknown prevalence

We analyze in this subsection how much of a reduction in the number of tests is achieved if the strategy, (3^*k*^, …, 3) with *k* = *k*_3_(*p*), is applied to a case with infection probability different from the actual *p*. The fact that *k*_3_(*p*), is a staircase function of *p* smears out the effect of the uncertainties on the value of *p*. As shown in Fig. 2, the intervals, (*p*_3,*k*+1_, *p*_3,*k*_], over which the strategy with *m* = (3^*k*^, …, 3) is best, span approximately half an order of magnitude. It is likely that the order of magnitude of *p* be known *a priori*. As it is apparent in Fig. 2, it is *p*_3,*k*_ and *p*_3,*k*+2_ that differ by approximately an order of magnitude. In fact, it can be deduced from Eq. (4) that (1 − *p*_3,*k*+2_)^9^ = (1 − *p*_3,*k*_) so that 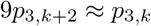. Thus, we will compare the use of a strategy with (3^*k*^, …, 3) for a situation with *p* ∈ (*p*_3,*k*+2_, *p*_3,*k*+1_) and of another with (3^*k*+1^, …, 3) for a situation with *p* ∈ (*p*_3,*k*+1_, *p*_3,*k*_). The first analysis will also shed light on how to handle cases in which the optimal *m*_1_ exceeds the maximum pool size allowed for the tests to give reliable results.

We show in Fig. 5 (a) plots of *D*_*k*_(*m, p*) *vs p* for strategies of the form *m* = (3^*k*^, …, 3) with *k* = 1 (black), *k* = 2 (red), *k* = 3 (green) and *k* = 4 (blue). It is apparent in this figure that the (*k* + 1)−th stage strategy is the best one for *p* ∈ (*p*_3,*k*+1_, *p*_3,*k*_) and that, at the transition probability, *p*_3,*k*+1_, it is *D*_*k*_ = *D*_*k*+1_ with *D*_*k*_ *< D*_*k*+1_ for *p > p*_3,*k*+1_ and *D*_*k*_ ≥ *D*_*k*+1_ otherwise. The figure also shows that even if not optimal, the (*k* + 1)-th stage strategy will produce a noticeable reduction in the number of tests per individual for *p < p*_3,*k*+1_. In fact, the discussion on Fig. 3 showed that when a strategy of the form *m* = (3^*k*^, …, 3) is applied to situations with *p* smaller than the smallest value for which the strategy is optimal (*i.e*., *p < p*_3,*k*+1_), initial pools with more than one infected sample become increasingly rare. In fact, if *p* is small enough the number of infected samples in a finite population will be almost zero. The expected number of tests per individual will be correctly approximated by Eq. (7) and, because of its low value, it will also be smaller than the standard deviation. This is illustrated in Fig. 6 (a) where we show the results of stochastic numerical simulations in which we apply the strategy with *m* = (3^*k*^, …, 3) and *k* = 3, which is optimal for *p* ∈∼ (0.01347, 0.03987), to a situation with infection prevalence, 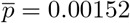, (41 infected samples over 27000 analyzed). As before, the figure shows the histogram (in log scale and as fraction of occurrences) of the number of tests performed per initial pool obtained with 1000 numerical realizations of the strategy (*i.e*., with 1000 initial pools of 27 samples each). We infer from the figure that, within these 1000 realizations, there are no instances in which the initial pool had more than one infected sample. The pools with exactly one infected sample are those requiring 10 tests to be resolved and the rest corresponds to pools with no infected samples and only one test. In this case the resulting average number of tests per individual, 0.0507, is equal to the one given by Eq. (7) with 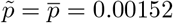 and the resulting standard deviation is 0.066. We contrast this distribution with the one that is obtained with the strategy, *m* = (3^*k*^, …, 3), that is optimal for *p* ≈ 0.0015. We show in Fig. 6 (b) the histogram obtained with 1000 realizations of this strategy for which it is *k* = 6 (729,000 samples analyzed, 1133 infected). In this case, even if there are pools that need many more than 10 tests to be resolved, starting with a larger number of samples (*m*_1_ = 749) results in a smaller average number of tests per individual, 0.0264, and standard deviation, 0.0212, than when using the suboptimal strategy with *k* = 3. If we restrict the number of realizations of the optimal strategy to 37, so that a similar number of individual samples (27,713) is analyzed as in the 1000 realizations of the strategy with *k* = 3 (27,000), a smaller number of tests per individual (average number = 0.0256 with standard deviation= 0.0218) is obtained compared to that of the suboptimal case. However, in the analyzed example, the suboptimal case still produces a very noticeable reduction in the number of tests. Even when the resulting standard deviation, 0.066, is slightly larger than the expected value, 0.0507, it is clear that practically no pool will require more than 10 tests to be resolved (which is *<* 40% of the number of samples in the initial pool). In general, if we apply the *m* = (*µ*^*k*^, …, *µ*) strategy to a situation with *p* ≪ *p*_*µ,k*+1_, not only the expected number of tests per individual, (1 + *kµ*)*/µ*^*k*^, will be given by Eq. (7), but more likely the number of tests per pool required will never exceed (1 + *kµ*), which is smaller than the number of individuals tested per initial pool, *m*_1_ = *µ*^*k*^ for *k* ≥ 2 and *µ* ≥ 3.

**Fig 5.**
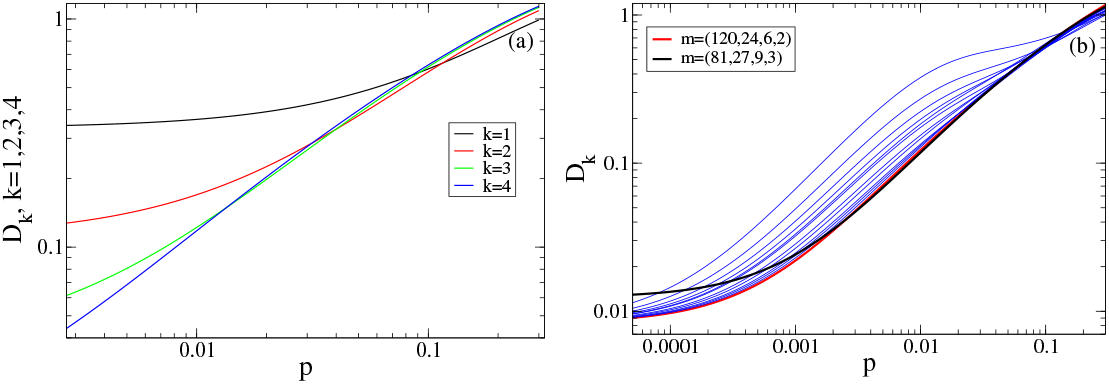
Cost of different nested strategies. (a) *D*_*k*_(*m, p*) *vs p* for strategies of the form *m* = (3^*k*^, …, 3) with *k* = 1 (black), *k* = 2 (red), *k* = 3 (green) and *k* = 4 (blue). (b) *D*_*k*_(*m, p*) *vs p* for the strategies with *m* = (120, 20, 4), *m* = (120, 15, 3), *m* = (120, 12, 3), *m* = (120, 10, 2), *m* = (120, 8, 2), *m* = (120, 6, 2), *m* = (120, 6), *m* = (120, 5), *m* = (120, 4), *m* = (120, 3) and (120, 2) (blue), *m* = (120, 24, 6, 2) (red) and *m* = (81, 27, 9, 3) (black).

**Fig 6.**
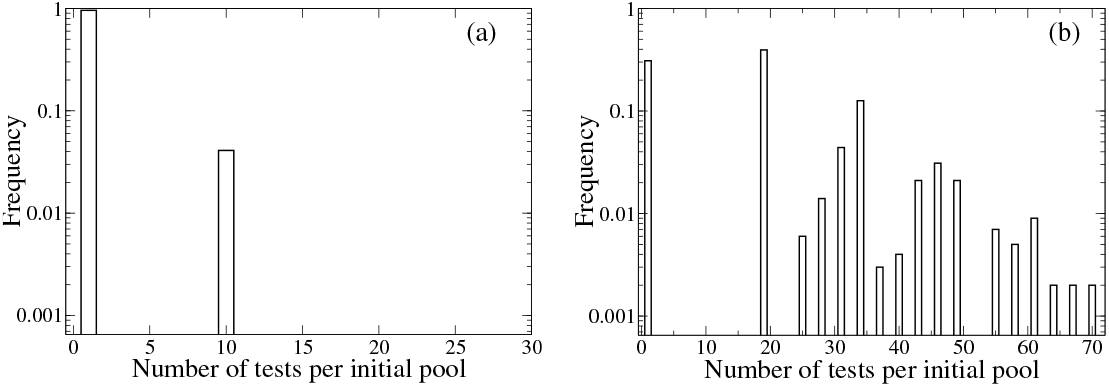
Number of tests of the nested strategy in optimal and sub-optimal conditions. Histograms (as fraction of occurrences in log scale) of the number of tests performed per pool derived from 1000 numerical realizations in which strategies of the form *m* = (3^*k*^, …, 3) are applied to a situation with infection prevalence, *p* = 0.00152, using a sub-optimal, *k* in (a) (*k* = 3, 27,000 individual samples analyzed, 41 infected) and the optimal in (b) (*k* = 6, 729,000 samples analyzed, 1133 infected).

*Based on the discussion of this Section we conclude that, if the value of p is poorly known a priori, it is advisable to use a strategy with m* = (3^*k*^, …, 3) *and a relatively small number of stages*, (*k* + 1). As discussed in S1 Appendix, Sec. 2, after the application of the first stage of the strategy to several pools, the estimate of *p* can be improved and the strategy (*i.e*., the value of *k*) be retuned.

### Maximum number of samples allowed in a pool

We now analyze what can be done if the optimal strategy requires a value, *m*_1_, that is larger than the maximum allowed for the test to be reliable. To perform this analysis we arbitraritly set the bound *m*_1_ ≤ 120. We then assume that we are dealing with a situation such that 0.00150588 *< p <* 0.00451083 for which the optimal strategy of the form (3^*k*^, …, 3) has *k* + 1 = 6 stages so that *m*_1_ = 3^5^ = 243 *>* 120. The demonstration of the theorem that solves the optimization problem in [14] involves a series of preliminary proofs, some which are useful to find a solution of the optimization problem with constraints. These proofs show that, for optimal solutions the ratio, *m*_*j*_*/m*_*j*+1_, cannot be smaller than the ratio, *m*_*j*+1_*/m*_*j*+2_ and that *m*_*j*_*/m*_*j*+1_ cannot be equal to 2 for two different values of *j*. In view of these results, any optimal strategy with constraints can only divide a given pool into 2 subpools at the very last stage (when going from a multi-sample pool to the individual sample testing). These two properties restrict the set of possible strategies over which to search for an optimal one when there is a constraint, for example, an upper bound on the number of samples in a pool. The search can be done algorithmically as explained in S1 Appendix, Sec. 1. In particular, if *m*_1_ = 120 the strategies whose costs should be compared to decide the optimal are: *m* = (120, 24, 6, 2), *m* = (120, 20, 4), *m* = (120, 15, 3), *m* = (120, 12, 3), *m* = (120, 10, 2), *m* = (120, 8, 2), *m* = (120, 6, 2), *m* = (120, 6), *m* = (120, 5), *m* = (120, 4), *m* = (120, 3) and (120, 2).

We compare in Fig. 5 (b) these 11 strategies that have *m*_1_ = 120 (all in blue but the one with *m* = (120, 24, 6, 2) in red) with the 5-stage strategy, *m* = (3^4^, 3^3^, 3^2^, 3) (black), which is optimal, according to the theory, for 0.00451083 *< p <* 0.0134716. We observe that the *m* = (3^4^, 3^3^, 3^2^, 3) scheme gives the largest reduction in the number of tests for 0.0025, :S *p* and that, even if not optimal, it does give a noticeable reduction for smaller values of *p*. For the smallest values of *p*, the best among all these strategies is the one with *m* = (120, 24, 6, 2). As *p* is reduced beyond the values illustrated in the figure, the cost of all the strategies that start with *m*_1_ = 120 approach the same value, which is smaller than the cost approached by the *m* = (3^4^, 3^3^, 3^2^, 3) strategy. Namely, when *p* becomes small enough (and there is at most one infected sample per pool at all stages) the expected number of tests per individual is given by Eq. (7) with 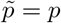 and is dominated by the term, 1*/m*_1_. Thus, the larger *m*_1_, the larger the reduction.

*We then conclude that, given an upper bound, m*_1,*max*_, *on the number of samples that can be combined in a pool, and an infection probability, p, such that* 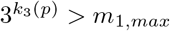 *it is advisable to use the strategy, m* = (3^*k*^, …, 3), *with k* = ⌊log_3_(*m*_1,*max*_) *⌋, if p is not much smaller than the probability for which the strategy is optimal. For very small p, larger reductions are achieved as m*_1_ *grows, regardless of whether m*_1_ = 3^*k*^ *or not. The search for the optimal strategy, on the other hand, can be done algorithmically as described in S1 Appendix, Sec. 1, taking advantage of various proofs of [14] that restrict the space over which the search has to be performed. Starting with m*_1_ = 3^*k*^, *however, has other advantages as discussed in the following*.

### Running the nested strategy in parallel

When running the *m* = (*m*_1_, …, *m*_*k*_) nested strategy in parallel for *N*_*w*_ pools (*i.e*., initially testing pooled samples of *N* = *N*_*w*_ *m*_1_ individuals), the expected number of tests performed at each stage, *j*, is 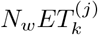 with 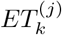 given by Eq. (2). These numbers are to be compared with *N*_*w*_ to design any practical implementation of the nested strategy. This is equivalent to comparing 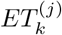 to 1. We show in Fig. 7 (a) a plot of 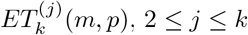, as a function of *p* for the strategies, *m* = (3^*k*^, …, 3), with *k* = 1 (black), *k* = 2 (red), *k* = 3 (green) and *k* = 4 (blue). The plotted curves are drawn with thicker lines for the range of *p* values for which each of the schemes is optimal. The horizontal line (magenta) indicates the number of tests that are performed per initial pool (always equal to 1). We observe that 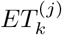(*m, p*) is an increasing function of *p* for every *j* and *k*. This means that within its range of optimal performance, 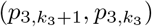, the scheme with 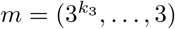 will require the largest number of tests (on average) for 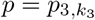. As shown by the figure, the number of tests that can be expected at the second stage (*j* = 2) is about twice as large as the number of tests of the first stage for 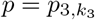 while it is almost equal for 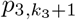. We also observe that the number of tests expected for the *j* − th stage increases with *j*. This means that we can expect the largest number of tests at the last stage, *k* + 1. The ratio of the number expected for two subsequent stages, however, never exceeds ∼ 2.1 for any, *p*, smaller than the maximum value for which the strategy is optimal. This is reflected in Fig. 7 (b) where we have plotted the ratio 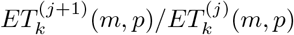 with *m* = (3^*k*^, …, 3) and 1 ≤ *j* ≤ *k* for *k* = 1 (black), *k* = 2 (red), *k* = 3 (green), *k* = 4 (blue) and *k* = 5 (magenta). We observe that all the ratios decrease with decreasing *p* and that, while those with *j* ≥ 2 approach 1 as *p* → 0, the ratio, 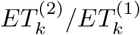, goes to zero (as 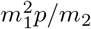). Interestingly, the ratios for *j* ≥ 2 are indistinguishable for any given *k*. Many of these features are reflected, for example, in the simulations of Fig. 3 where the average numbers of tests performed per initial pool at the second, third and fourth stages are, respectively: 1.284, 1.545, 1.605 (a); 2.001, 2.808, 3.099 (b) and 0.945, 1.068, 1.092 (c).

**Fig 7.**
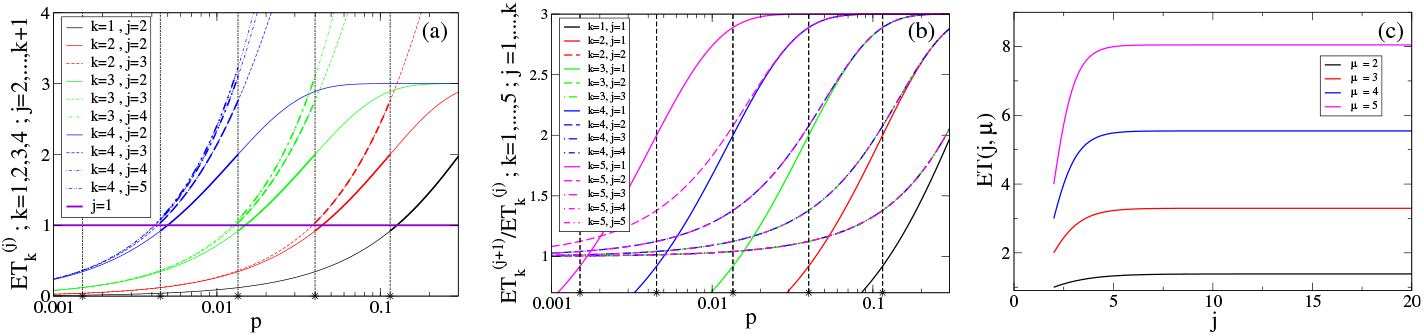
Expected number of tests per stage. (a) Expected number of tests, 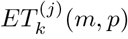, at each stage, *j*, with 2 ≤ *j* ≤ *k*, as a function of *p* for strategies of the form *m* = (3^*k*^, …, 3) with *k* = 1 (black), *k* = 2 (red), *k* = 3 (green) and *k* = 4 (blue). The thicker portion of the curves correspond to the values of *p* for which each of the schemes is optimal. The horizontal line in magenta corresponds to the number of tests at the first stage (*j* = 1) for all *k*. (b) Ratio of the number of tests expected for two subsequent stages, 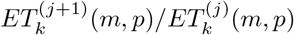, of the strategies with *m* = (3^*k*^, …, 3) and *k* = 1, .., 5. (c) 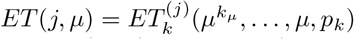, as a function of *j* for *µ* = 2 (black), *µ* = 3 (red), *µ* = 4 (blue) and *µ* = 5 (magenta). We plotted the curves for any value of *j*, but it must be noted that only the integer ones are meaningful.

Given that 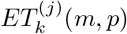 for *m* = (*µ*^*k*^, …, *µ*) decreases with *p*, it is meaningful to compute it for *k* = *k*_*µ*_ at *p* = *p*_*µ,k*_*µ* (*i.e*., at the maximum probability for which the scheme 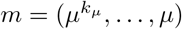 is optimal). Thus:

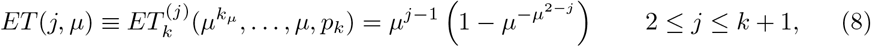

which is independent of *k*. We show in Fig. 7 (b) plots of *ET* (*j, µ*) as functions of *j* (allowing *j* to take any real number) for various values of *µ*. We observe that *ET* (*j, µ*) approaches a limiting value, **ET**_*µ*_, with increasing *j*. This limiting value, which puts an upper bound on the number of tests that one can expect to be performed at any stage starting from a pool with 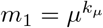 samples using a scheme of the form *m* = (*µ*^*k*^, …, *µ*) for a population with *p* ≤ *p*_*µ,k*_*µ*, is

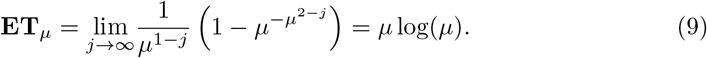

This implies that, if one uses a scheme of the form 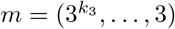 for situations in which the infection probability is 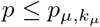 then we can expect that the number of tests at each stage will never be above **ET**_**3**_ ∼ 3.296 times the number of tests of the first stage. The upper bound, **ET**_*µ*_, increases with *µ*. This means that the largest *µ* the largest will be the expected number of tests at any given stage compared with the first one (provided that *p* is smaller than the corresponding *p*_*µ,k*_). This is an important feature when one wants to run the proposed scheme on several first stage pools in parallel and there is a limit on the number of tests that can be performed simultaneously. At the first stage one would have a number of pools equal to this limit. Thus, a less frequent overflow of tests with respect to the first stage will facilitate the bookkeeping of the method.

We show in Tables 1 and 2 the results of stochastic numerical simulations performed as before but this time assuming that at most 96 tests can be run simultaneously at any given stage and that at most 32 individual samples can be mixed in any given pool. The upper bound on the number of simultaneous tests comes from one of the typical sizes of commercially available multi-well plates where to run the tests. On the other hand, the maximum number of individual samples that can be mixed to detect the presence of a single one infected with SARS-CoV-2 using RT-qPCR has been estimated to be 32 in [30]. Table 1 corresponds to a situation in which the infected population is *p* = 0.02 for which the optimal number of stages for a scheme of the form (3^*k*^, …, 3) is *k* = *k*_3_ = 3. We then compare in this table the results of running the tests in parallel for the (3^3^, 3^2^, 3) scheme and for a scheme that starts with *m*_1_ = 32. We use the same population in both cases, with some added samples in the latter case. While the former scheme is optimal for *p* = 0.02 and satisfies that *m*_1_ *<* 32, the latter puts the largest number of allowed individual samples in each of the first pools. Even if the former starts with fewer samples than the second one, it allows to resolve more cases after the first parallel run because fewer pools (and samples) are left for the second round of stages. Furthermore, the number of tests per individual turns out to be smaller for the (3^3^, 3^2^, 3) scheme. Changing the values of *m*_2_ and *m*_3_ in the scheme that starts with *m*_1_ = 32 does not improve its performance. Namely, the case with *m*_2_ = 16 and *m*_3_ = 4 does not resolve all cases in two parallel rounds, but needs a third one. This is consistent with the proofs of [14] that we referred to before. We then perform the same comparison for *p* = 0.008 for which the optimal scheme of the form (3^*k*^, …, 3) has *k* = *k*_3_ = 4. In this case we would have to start with *m*_1_ = 2^4^ = 81 samples which is not allowed. Then, we compare the same two schemes as before and a scheme that starts with *m*_1_ = 32 but has *k* = 4 instead of *k* = 3. We show the results in Table 2. There we see that there is no pool overflow in this case, something that we could have expected for the (3^*k*^, …, 3) scheme based on the discussion of Fig. 7. The comparison shows that the (3^*k*^, …, 3) scheme is the one that requires the smallest number of tests per individual even if the number of stages is not the optimal for the infection probability of the population.

**Table 1.** Comparison between the optimal strategy and one that starts with the maximum allowed number of samples in a pool.

| **$m_1 = 27, m_2 = 9, m_3 = 3, m_4 = 1$. Individuals tested: 2592. Infected samples: 53.** |  |  |  |  |  |
| --- | --- | --- | --- | --- | --- |
| Stage | Tests | Infected pools detected | Pool overflow | Not-infected samples identified | Infected samples identified |
| j=1 | 96 | 39 | 0 | 1539 | 0 |
| j=2 | 96 | 41 | 21 | 495 | 0 |
| j=3 | 96 | 35 | 27 | 183 | 0 |
| j=4 | 96 | 32 | 9 | 64 | 32 |
| Accumulated Number of Cases Informed |  |  |  | **2281** | **32** |
| j=2' | 21 | 8 | 0 | 117 | 0 |
| j=3' | 51 | 18 | 0 | 99 | 0 |
| j=4' | 63 | 21 | 0 | 42 | 21 |
| Accumulated Number of Cases Informed |  |  |  | **2539** | **53** |
| **Tests/individual = 0.2002** |  |  |  |  |  |
| **$m_1 = 32, m_2 = 8, m_3 = 4, m_4 = 1$. Individuals tested: 3072. Infected samples: 64.** |  |  |  |  |  |
| Stage | Tests | Infected pools detected | Pool overflow | Not-infected samples identified | Infected samples identified |
| j=1 | 96 | 48 | 0 | 1536 | 0 |
| j=2 | 96 | 29 | 96 | 536 | 0 |
| j=3 | 58 | 30 | 0 | 112 | 0 |
| j=4 | 96 | 25 | 24 | 71 | 0 |
| Accumulated Number of Cases Informed |  |  |  | **2255** | **25** |
| j=2' | 96 | 31 | 0 | 520 | 0 |
| j=3' | 62 | 32 | 0 | 120 | 0 |
| j=4' | 96 | 25 | 56 | 71 | 25 |
| Accumulated Number of Cases Informed |  |  |  | **2966** | **50** |
| j=4'' | 56 | 14 | 0 | 42 | 14 |
| Accumulated Number of Cases Informed |  |  |  | **3008** | **64** |
| **Tests/individual = 0.2135** |  |  |  |  |  |

Results of stochastic numerical simulations in which the nested pool strategy is performed simultaneously on up to 96 pools each of which can have at most 32 samples. At the first stage there are exactly 96 pools, but at the second stage the number can be larger. This is what we call “pool overflow” in the table. These unresolved pools can be tested whenever there is room. In the table we put in the same row all the pools of equal size that are available for testing at that point (identifying them with their corresponding value of $j$ ). The optimal strategy for $p = 0.02$ (the one with $m_1 = 27$ ) produces less overflow than those that start with the largest allowable number of samples in a pool ( $m_1 = 32$ ).

**Table 2.** Comparison of strategies when the optimal includes pool sizes that are too large for a reliable detection.

| **$m_1 = 27, m_2 = 9, m_3 = 3, m_4 = 1$. Individuals tested: 2592. Infected samples: 19.** |  |  |  |  |  |
| --- | --- | --- | --- | --- | --- |
| Stage | Tests | Infected pools detected | Pool overflow | Not-infected samples identified | Infected samples identified |
| j=1 | 96 | 15 | 0 | 2187 | 0 |
| j=2 | 45 | 19 | 0 | 234 | 0 |
| j=3 | 57 | 19 | 0 | 114 | 0 |
| j=4 | 57 | 19 | 0 | 38 | 19 |
| Accumulated Number of Cases Informed |  |  |  | **2573** | **19** |
| **Tests/individual = 0.0983** |  |  |  |  |  |
| **$m_1 = 32, m_2 = 8, m_3 = 4, m_4 = 1$. Individuals tested: 3072. Infected samples: 25.** |  |  |  |  |  |
| Stage | Tests | Infected pools detected | Pool overflow | Not-infected samples identified | Infected samples identified |
| j=1 | 96 | 21 | 0 | 2400 | 0 |
| j=2 | 84 | 24 | 0 | 480 | 0 |
| j=3 | 48 | 24 | 0 | 96 | 0 |
| j=4 | 96 | 25 | 0 | 71 | 25 |
| Accumulated Number of Cases Informed |  |  |  | **3047** | **25** |
| **Tests/individual = 0.1055** |  |  |  |  |  |
| **$m_1 = 32, m_2 = 16, m_3 = 8, m_4 = 4, m_5 = 1$. Individuals tested: 3072. Infected samples: 25.** |  |  |  |  |  |
| Stage | Tests | Infected pools detected | Pool overflow | Not-infected samples identified | Infected samples identified |
| j=1 | 96 | 21 | 0 | 2400 | 0 |
| j=2 | 42 | 22 | 0 | 320 | 0 |
| j=3 | 44 | 24 | 0 | 160 | 0 |
| j=4 | 48 | 24 | 0 | 96 | 25 |
| j=5 | 96 | 25 | 0 | 71 | 25 |
| Accumulated Number of Cases Informed |  |  |  | **3047** | **25** |
| **Tests/individual = 0.1061** |  |  |  |  |  |

*Based on the above discussion, we conclude that the schemes with m* = (3^*k*^, …, 3) *are not only good because of the relatively fewer number of tests per individual they require (even for probabilities for which they are not optimal), but also because they produce some of the smallest overflows when running the strategy in parallel. In clinical laboratories the turnaround time directly impacts on the productivity. Thus, even when strategies in which the pools are subsequently divided in two subpools produce less overflow, they require more stages and, consequently, more time to identify the infected samples than those in which the pools are subdivided in three*.

### Taking advantage of the nested strategy for result verification and self-consistency tests

Any test carries some level of uncertainty. There are two main ways to reduce this uncertainty. One way is to employ novel technologies, to improve the limit of detection and therefore the test sensitivity, and use commercial kits less prone to interference. The other way is to repeat the test several times. A nested strategy as the one described in this paper has the advantage that a fraction of the samples is retested several times, particularly those that are identified as infected. The quantification of the analyte, on the other hand, could provide self-consistency checks to increase the reliability of the results. Droplet Digital PCR (ddPCR) is ideally suited both to improve the sensitivity of the detection and to quantify the detected material. It has been observed, particularly for the case of SARS-Cov-2, that ddPCR can detect RNA contents between one and two orders of magnitude below what RT-qPCR detects [23, 25, 31]. ddPCR is an end-point method that quantifies the nucleic acid content (provided that it is within a certain range) without the need of calibration curves [32, 33] (S1 Appendix, Sec. 3 for details).

The increased sensitivity of ddPCR also enhances the reliability of the detection of the reporters that are used in any PCR test to confirm that the reaction has proceeded correclty as well as the integrity of the sample, avoiding false negative results due to inappropriate handling. As an internal control, a human RNA molecule expected to be found in all the samples regardless of the presence of viral RNA is co-amplified. If the control RNA is not detected, it is assumed tnat the sample was inappropriately obtained or handled or a reaction inhibitor is present. In such a case, the result of the test is not reported and a new sample from the individual is requested. When multiple samples are pooled, the human RNA contained in only one original sample can provide the signal indicating the presence of the human RNA, masking tha absence of such RNA in the remaining samples of the pool. The only negative results that could be informed reliably would be those tested at the level of individual samples. In the case of our strategy, this would occur at the very last stage, a stage that is reached by relatively few non-infected samples compared to those contained in pools that do not test positive at previous stages. The fraction of non-infected samples, on the other hand, should be at least two thirds of those analyzed for the application of pool testing to make sense (see Eq. (1)). Not informing most of the non-infected samples means that the strategy would be applied mainly to identify those that are detectably infected. In other words, it could be technically possible that all the samples for which both, no signal of viral RNA is detected and have not been tested individually in the process of pool partitioning, are false negatives. This type of events have been shown to be very unlikely and of having a larger probability of false negatives is currently accepted as a limitation of the pooling method with no significant clinical meaning. Considering these limitations, test pooling may still serve for epidemiological purposes and for continuous validation of the method. As analyzed in this Section, however, accurate quantification that ddPCR provides can be of help to enlarge the truly confirmed set of negative tests.

We discuss in what follows how a self-consistency check could be applied to the results obtained with our strategy when the nucleic acid content is quantified as described in S1 Appendix, Sec. 3, and how this quantification can be used to detect some of the flawed pooled samples that test negative. We also use the ddPCR quantification to determine the probability of detecting a single infected sample in a pool as a function of the pool size and the viral load.

### Test verification

In ddPCR the volume that goes in the reaction tube is subdivided into many (20,000) sub-volumes. At the end, the test gives (ideally) the number of sub-volumes that contained, at the beginning of the test, at least one molecule of the nucleic acid of interest (RNA in our case). As explained in S1 Appendix, Sec. 3, if the fraction of occupied sub-volumes is bounded away from 0 or 1, a range of possible values for the concentration of the RNA detected by the test can be obtained. As derived in S1 Appendix, Sec. 3, when testing a pool of *m*_*j*_ samples, the RNA concentration that is estimated at the end of the ddPCR corresponds to:

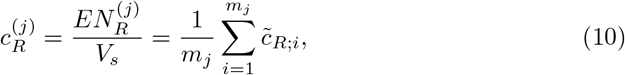

where the indices, *i* = 1, …, *m*_*j*_, identify the samples contained in the pool, 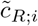 is the RNA concentration of the *i*-th sample extracted from the corresponding individual (what we call the *original sample* in S1 Appendix, Sec. 3) and 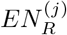 is the expected number of RNA molecules in a volume of the pool equal to the *sample volume, V*_*s*_. *V*_*s*_ does not involve any dilution with respect to the original sample exctracted from the individual. The preparation for the test can be accounted for as introducing a dilution of factor, *D*, that does not change the number of RNA molecules of interest contained in *V*_*s*_; *i.e*., 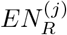 is also the number of RNA molecules that we expect to find for the given pool in the volume of the test, *V*_*t*_ = *DV*_*s*_. Under the usual assumptions for the quantification in ddPCR tests, it will possible to estimate the concentration provided that it satisfies 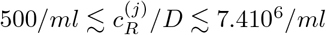 (see S1 Appendix, Sec. 3, where we also discuss how this range could eventually be enlarged).

We discuss now how the nested nature of the strategy can be used for test verification. Let us then consider an *m*_*j*_-sample pool, *m*_*j*_ *>* 1, that tests positive at the *j*-th stage so that, at stage *j* + 1, is sub-divided into *m*_*j*_*/m*_*j*+1_ sub-pools with *m*_*j*+1_ samples in a sample (test) volume, *V*_*s*_ (*V*_*t*_), each. In view of Eq. (10), the expected number of RNA molecules added over all the (*m*_*j*_*/m*_*j*+1_) *m*_*j*+1_-sample sub-pools of the *m*_*j*_-sample pool is:

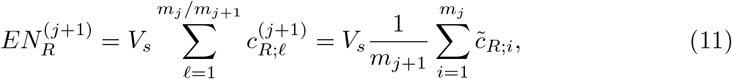

where 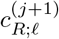 is the RNA concentration, before dilution, that is estimated for the, ℓ-th sub-pool at the end of the ddPCR test of the *j* + 1-th stage (see S1 Appendix, Sec 3 for more details). Eqs. (10) and (11) imply that:

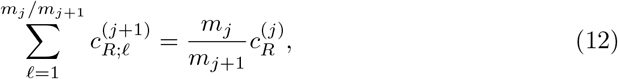

which is a condition that the RNA concentrations derived from the ddPCR tests performed on the same subset of samples at two subsequent stages should satisfy. As explained in S1 Appendix, Sec. 3, the ddPCR quantification gives an interval of possible values for the corresponding concentration, *c*_*R*_. The uncertainty, however, can be relatively low compared with the expected value, especially away from the lowest end of concentrations for which the quantification is possible (see Fig. 3 in S1 Appendix). In the case of low viral load, the RNA content could be very small, especially for large pools. In the case of the human RNA it is possible that the upper bound be reached. This can be avoided by choosing judiciously the piece of human RNA to be detected. Depending on the infection, the upper bound might also be reached for the viral RNA concentration in the case of individual samples. In the case of individuals infected with SARS-CoV-2, for example, the viral RNA detected could be larger than ∼ 10^7^ copies/*ml* [34, 35] in throat samples (even in the testing volume) and reach ∼10^11^ copies/*ml* in sputum [36]. In any case, the comparison of the RNA concentrations derived in two subsequent stages, either viral and/or human, can be used to verify that both stages have proceeded as expected. The advantage of the nested strategy is that the set of samples that would require some re-testing, if an inconsistency is found, is reduced with respect to strategies that are either non-adaptive or that mix up the samples at random at every stage.

Knowing the “genealogy” of the infected samples, *i.e*., the pools they belonged to at any given stage, is also of help for the quantification of the viral RNA content when its concentration in the infected individual sample is too high. Namely, if at the last stage this concentration exceeds the upper bound allowed for a reliable quantification (see S1 Appendix, Sec. 3), then, in certain cases it is possible to deduce it from the concentration determined at a previous stage.

Let us consider the example of Fig. 1, which corresponds to a pool, *W*_0_, with 9 samples at the initial stage, only one of which is infected. The infected sample, *W*_020_, is identified at the very last stage. Let us use the same labeling that we use for pools and samples to identify the RNA concentrations in each of them and let us assume that 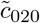 is large enough so that 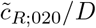 exceeds the upper bound for a reliable quantification with ddPCR (∼ 7.4 10^6^*ml*^−1^, see S1 Appendix, Sec. 3). Knowing the genealogy allows us to infer that this is the only infected sample in the second-stage pool, *W*_02_ and in the first-stage one, *W*_0_. In either of these two cases the RNA concentration is diluted with respect to the last stage (by factors 3 and 9, respectively). Therefore, it is possible that the RNA viral concentration in *W*_02_ or in *W*_0_ could be quantified. Since we know that it is only due to the (020) sample, we can then infer, 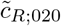. Namely, the application of Eq. (10) to the pools that contain the (020) sample at stages *j* = 1 and *j* = 2 implies that

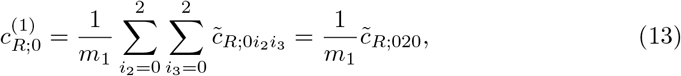

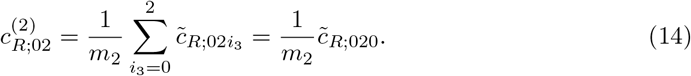

Clearly, 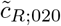 can be estimated using either Eq. (13) or (14), depending on whether 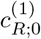 or 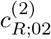 can be quantified with ddPCR. Furthermore, the estimate can be computed as an average of the values obtained at the two stages reducing its uncertainty in this way. This same reasoning can be applied to the example of Fig. 1, S1 Appendix, Sec. 1, where there are more options because there are more stages. In the case of the samples, (0112) and (0122), that remain in the same pool at stage *j* = 2, in particular, their viral load could be estimated provided that the RNA content of one of them could be quantified at stage *j* = 3.

### Minimum viral load detectability and maximum pool size using ddPCR

One of the main limitations of test pooling within the problem addressed in this paper is the loss of sensitivity due to the dilution of the samples in the pool. In that regard, the worst case scenario is that in which all the samples but one correspond to healthy individuals. Let us consider an *m*_*j*_-sample pool in which the *i*-th sample corresponds to an infected individual whose viral RNA concentration is 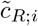 (defined as in Eq. (10)). Assuming that ddPCR allows the detection of up to a single RNA molecule in the testing volume, we compute the probability of having at least one viral RNA molecule in the pool, 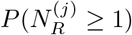, as a function of 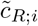 as (S1 Appendix, Sec. 4):

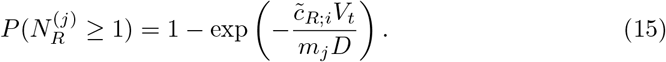

We show in Fig. 8 the plot of 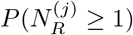 as a function of 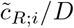 for *m*_*j*_ = 3 (black), 9 (red) and 27 (green), where we have used the testing volume, *V*_*t*_ = 20*µl*. The horizontal line corresponds to *P* = 0.99. The vertical lines correspond to the SARS-CoV-2 RNA content detected and quantified by ddPCR in the test volume of individual samples reported in [34]: the curve in blue corresponds to the minimal load detected (11.1 copies/test) and the one in purple to the average minus SEM (651-501 copies/test) detected in samples of nasal swabs (the specimens with lowest viral content reported in [34]). There we observe that a single sample with the minimal viral load detected in [34] could be detected, according to our calculation, in *>* 97% of the cases in 3-sample pools and in *>* 70% of the cases in 9-sample ones. The lowest end of the average obtained in nasal swabs, on the other hand, could be detected with 3, 9 or 27-sample pools in over 99% of the cases. Considering that the average viral loads detected in throat swabs and sputum samples were, respectively, ∼ 4 and 27 times the average in nasal swabs, it seems that a single infected sample would be detected in a fairly large fraction of cases using pools of 27 or more samples of these specimens.

**Fig 8.**
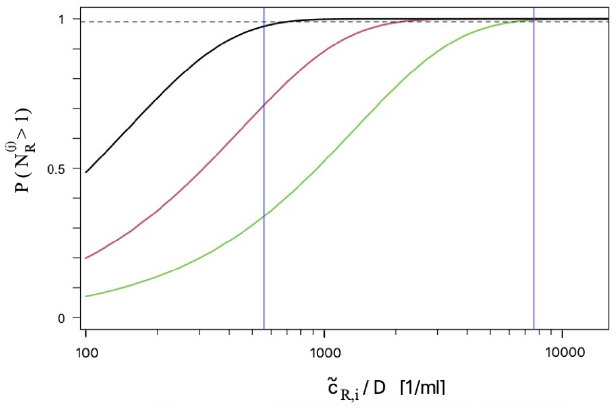
Probability of having at least one RNA molecule in the testing volume. We plot this probability for an *m*_*j*_ pool (black: *m*_*j*_ = 3, red: *m*_*j*_ = 9, green: *m*_*j*_ = 27) in which only one sample corresponds to an infected individual whose RNA viral concentration is 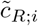. The blue vertical line corresponds to the minimal viral RNA amount (quantified with ddPCR) detected in individual samples of people infected with SARS-CoV-2 and the one in purple to the average minus SEM (651-501 copies/test) detected in nasal swabs [34]). The dashed horizontal line corresponds to the probability *P* = 0.99.

### Detecting flawed pooled samples

The aim of this analysis is to determine whether it is possible to identify “flawed” samples in pools that test negative. Here by “flawed” we mean samples with undetectable levels of the human RNA that should be present in any sample. Assuming that there is not a huge variability of its concentration among individuals we can expect that pools of all stages should be characterized approximately by the same concentration, 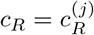, 1 ≤ *j* ≤ *k* + 1, if the original samples are not flawed and the PCR tests proceed correctly. Now, not only the quantification with ddPCR has an uncertainty but also the human RNA content varies between individuals. The question is how the variability in the values, *c*_*R*_, that may be derived with ddPCR compare with the change that one flawed sample in a pool would induce in *c*_*R*_. To analyze this situation, we introduce the notation

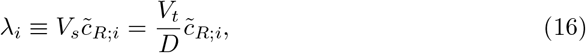

to denote the *mean number* of human RNA molecules of the *i*-th individual in a volume of size, *V*_*s*_. Let us then consider an *m*-sample pool in which each individual sample occupies a volume, *V*_*s*_*/m*, of the sample volume, *V*_*s*_. In such a case the *i*-th individual contributes with a number of human RNA molecules of mean *λ*_*i*_*/m*. Including the human to human variability implies that the values, *λ*_*i*_, are (independent) instances of a random variable with a certain distribution (if none of the individual samples are flawed). This is a distribution at the level of the human population that, for simplicity, we consider it is Normal of mean, *λ*, and standard deviation, *σ* (see S1 Appendix, Sec. 5). We then consider that an *m*-sample pool in which none of the samples is flawed is characterized by a sequence, *λ*_1_, *λ*_2_, …, *λ*_*m*_ of independent identically (Normal) distributed (i.i.d.) random variables. If there is one flawed sample in the pool, then, the corresponding *λ*_*i*_ will be zero and the other *m* − 1 will be i.i.d. random variables.

We now consider a ddPCR experiment in which the testing volume is divided into *M* sub-volumes. Proceeding as explained in S1 Appendix, Sec. 5, we compute the likelihood that *x* of the *M* sub-volumes come out as not having any RNA molecules at the end of the test under the two situations that we want to distinguish: (i) all the individual samples were “good”, *i.e*., they all contributed with human RNA molecules to the original sample volume, *V*_*s*_; (ii) one of the samples was “flawed”, *i.e*. did not contribute any human RNA to this volume. We obtain:

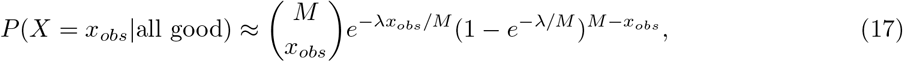

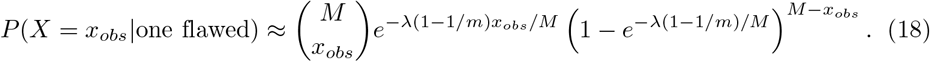

Therefore, the accuracy of the test (the fraction of instances for which the classification is correct) can be written as:

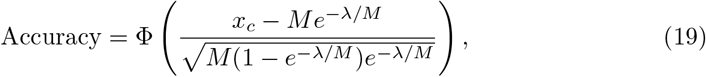

where 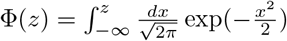 and the threshold, *x*_*c*_, is given by:

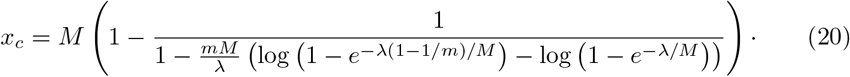

For very small and very high *λ* values Eq. (19) is not good. Namely, in such cases the distributions of the number of sub-volumes with no human RNA are concentrated around zero (for large *λ*) or *M* (for small *λ*) where the normal approximation that is used in Eq. (19) fails. Nevertheless, for those values of *λ* the accuracy tends to 1/2. We show in Fig. 9 a plot of the accuracy given by Eq. (19) as a function of *λ*, the mean number of human RNA molecules in a volume, *V*_*s*_, of the original sample for 3 pool sizes (*m* = 3, 9, 27) and *M* = 20, 000, a standard for ddPCR tests. We observe in the figure that, for the accuracy to be greater than 0.99 (dotted line) *λ* can vary within the interval, [172, 2.96 10^5^], if the pool size is *m* = 3. In the *m* = 9 case, it must satisfy 1.7 10^3^ ≤ *λ* ≤ 1.36 10^5^. For a pool with *m* = 27 we can not obtain an accuracy of 0.99 for any value of *λ*. The general rule is that, if *m* is below a certain value, *m*_*max*_, there is a finite range of *λ* values over which the accuracy is larger than 0.99. This range gets narrower as *m* increases. If *m > m*_*max*_ the accuracy is below 0.99 for any value of *λ*.

**Fig 9.**
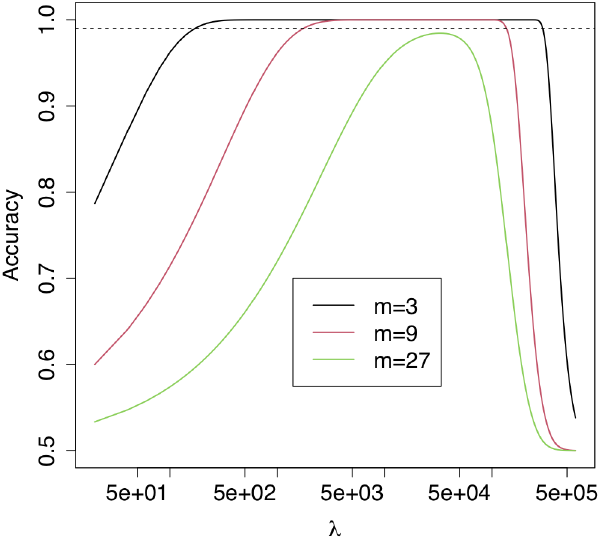
Statistical test to detect a flawed sample. We plot the accuracy of the test that discriminates between having one flawed sample in an *m*-sample pool and having none as a function of the expected number of RNA molecules, *λ*, in a volume of size, *V*_*s*_, of the original sample, when a ddPCR is run on the pool subdividing the testing volume, *V*_*t*_ = *DV*_*s*_, into *M* = 20, 000 sub-volumes. The pool sizes illustrated are *m* = 3 (black), *m* = 9 (red) or *m* = 27 (green). Here *λ* is the expected number over the whole human population. The dotted line corresponds to the accuracy, 0.99.

The range of *λ* values for which the discrimination test has high accuracy may be translated into RNA concentrations, *c*_*R*_, as done in Eq. (16), *i.e*., *c*_*R*_ = *λ/V*_*s*_ = *Dλ/V*_*t*_.

Using *V*_*t*_ = 20*µl* (a typical testing volume) we obtain that the concentrations in the orginal sample, *c*_*R*_, must satisfy 8.6 10^3^*/ml < c*_*R*_*/D <* 1.48 10^7^*/ml* for *m* = 3 and 8.5 10^4^*/ml < c*_*R*_*/D <* 6.8 10^6^*/ml* for *m* = 9. The range in the *m* = 9 case is fully contained within the range for which ddPCR can provide an accurate estimation of the RNA concentration (see S1 Appendix, Sec. 3). The highest end for the *m* = 3 case goes beyond the limit set by ddPCR; their overlap occurs for:

8.6 10^3^*/ml < c*_*R*_*/D <* 7.4 10^6^*/ml*. Assessing whether these conditions can be satisfied in actual situations requires further studies. In case that this discrimination could be possible for two successive stages, a re-checking along the lines already discussed for the viral RNA could be performed. On the other hand, knowing the genealogy of the pools that test positive at the *k*-th stage (the ones containing the samples that are tested individually at the last stage), we can go backwards and identify sets of negative pools that might contain missing samples. This would give a further validation on top of what could be determined at the time that the corresponding pools test negative.

## Conclusions

In this paper we have analyzed in detail the adaptive nested strategy for test pooling introduced in [14] which is an extension of Dorfman’s algorithm for more than two stages. We have mostly focused, in particular, on its practical implementation to identify individuals infected with a virus whose RNA is detected with the test. The strategy is characterized by the number of stages, *k* + 1, and the sequence of pool sizes, *m* = (*m*_1_, …, *m*_*k*_), that satisfy *m*_*j*_*/m*_*j*+1_ ∈ ℕ_*>*1_ for 1 ≤ *j* ≤ *k* and *m*_*k*+1_ = 1. We have analytic expressions for its cost (the expected number of tests per individual) and optimal parameters (*k* and *m* that minimize the cost) as functions of the infection probability, *p* (S1 Appendix, Sec. 1). As proved in [14], it only makes sense to apply it if *p* satisfies Eq. (1). The cost scales as *p* log(1*/p*), implying a similar reduction as other strategies [13, 18], some of which have the disadvantage, for practical implementations, of requiring a number of stages that is unknown *a priori*. Given that the optimal within our approach is determined analytically, the time it will take for it to resolve the number of infected individuals within a certain population can be estimated from the very beginning. The analyses of the present paper have shown that it is advisable to use, whenever possible, a strategy with *m* = (3^*k*^, …, 3) and *k* given by Eq. (4) (*µ* = 3) (Fig. 2), even if there are some small *p* intervals for which the optimal strategy is of the form, *m* = (4 3^*k*−1^, …, 3).

In the present paper we have first analyzed the variability of the number of tests of the strategy around its expected value, which determines the cost studied in [14]. We found that the relatively large standard deviation is mostly due to the large number of pools that initially turn out to be negative. Thus, we can expect that fluctuations about the mean will more likely result in fewer tests per individual than the cost. We have then studied the impact of restrictions for the use of the strategy of the form, *m* = (3^*k*^, …, 3), that is optimal for the given infection probability, *p*. In particular, we have analyzed how to proceed when *p* is unknown *a priori* or when the optimal starting pool size is larger than the maximum allowed by the test for a reliable detection of the RNA of interest. We have shown that, if the number of stages, *k* + 1, is small enough, noticeable reductions in the cost are obtained even if the strategy is not optimal for the infection probability of the analyzed population (Fig. 5 (a)). We have also discussed how to estimate *p* before or during the application of the strategy (S1 Appendix, Sec. 2). Given an upper bound on the maximum pool size, depending on *p*, strategies that start with *m*_1_ ≠ 3^*k*^ could eventually reduce the cost more than those of the form, *m* = (3^*k*^, …, 3) (Fig. 5 (b)). In such a case the sequence of pool sizes, *m*, can be chosen algorithmically as explained in S1 Appendix, Sec. 1. The use of the strategies with *m* = (3^*k*^, …, 3) have other advantages that we observed when applying our method in parallel on many pools at the same time as would be done when using multi-well plates to run the PCR test. In particular, the ratio between the expected number of tests at any given stage of the strategy and the number performed at the first stage increases with the ratio of subsequent pool sizes (Fig. 7). This means that, if we perform the first stage test on *N*_*w*_ pools simultaneously (*N*_*w*_ is, *e.g*., the number of wells of the plate), at subsequent stages we will need more than *N*_*w*_ wells to accommodate the pools that come from those that tested positive at the first stage and that the number of additional wells will be larger, the larger the fraction between successive pool sizes of the strategy. Reducing this *overflow* of tests is good for bookkeeping purposes. The *m* = (3^*k*^, …, 3) strategies present an appealing good balance between cost reduction even under non-optimal conditions and relatively small overflows (Tables 1 and 2).

The use of pool testing for the detection of viral RNA allows a widespread surveillance of the population that might be infected keeping costs under control at the same time. However, it carries two main problems. One is related to detectability: what is the maximum number of samples that can be mixed together so that the viral RNA content coming from even a single infected sample can be detected? The other one is related to detecting flawed samples. When samples are tested individually, the quality is assured by detecting a specific human RNA that is co-extrated with the viral genome. In the absence of this detection, the result of the test is not informed. In this paper we have analyzed this problem assuming that ddPCR not only has a greater sensitivity with respect to RTqPCR for viral RNA detection, but it also allows for the direct quantification of the nucleic acid content in a large dynamic range (S1 Appendix, Sec. 3). Given that the present work was originally motivated by the current COVID-19 pandemic, we contrasted some of our results with data obtained from people infected with SARS-CoV-2. We have shown that, assuming that ddPCR allows the detection of a single RNA molecule in the tested volume, even the samples of lowest viral load reported in [34] could be detected in 3-sample pools in over 95% of the cases (Fig. 8). Samples with average viral RNA content, on the other hand, could be detected, according to our calculations, in pools of 27 samples or more. The problem associated to sensitivity loss due to pooling can be partially resolved if the samples are tested more than once. Our strategy does so with those that belong to pools that test positive at the first stage, which gives reassurance to those results. In this regard, the nested nature of our strategy is advantageous, especially along with the quantification of the RNA content. We have shown, in particular, how the quantification achieved at the various stages of the strategy could be used for self-consistency checks or to quantify the viral RNA content in samples with very high load for which the individual testing could not give such an estimate.

Regarding the detectability of unviable samples with inhibitors or degraded RNA (invalid samples) in pool testing, we analyzed the possibility of discerning between pools with *m* valid samples samples and pools with one invalid and *m* − 1 viable specimens. Assuming that the variability of the human RNA content among individuals was described by a Normal distribution, we found ranges of concentrations for which such discrimination was possible with more than 99% accuracy for *m* = 3 and *m* = 9 and with more than 95% accuracy for *m* = 27. The current standard is to use human RNase P RNA. Whether the corresponding RNA content can lie within the dynamic ranges established by our calculations in the testing volume needs further studies. In any case, if the analysis turns out to be positive, at least, for certain pool sizes, together with the nested nature of our strategy that allows to infer information on previous stages from the output of subsequent ones, it could open up the possibility of enlarging the scope of the use of pools for diagnostic purposes.

In summary, we have analyzed in detail a nested test pooling strategy, providing a clear guidance to establish the number of samples in each pool and how to partition the positive pools for the identification of infected individuals. The simplicity of the strategy, its easily parallelization and the advantages of its nested nature make it a very valuable option to faithfully detect viral RNA in a cost effective manner.

## Supporting information

Supplementary Calculations

## Data Availability

We will make the programs we wrote available at a website we are setting up (https://wp.df.uba.ar/pooling)

## Supporting information

### S1 Appendix. Auxiliary calculations

This Appendix has 6 Sections. Sec. 1 includes a detailed description of our nested strategy together with explanations of how the main analytic formulas that we use in the paper are derived. In Sec. 2 we show how the infection probability can be computed within the framework of our nested strategy. In Sec. 3 we show how the nucleic acid content is estimated in ddPCR experiments, determine the range of concentrations for which the quantification is reliable and present the calculations that are needed for the self-consistency tests that we discuss in the paper. In Sec.4 we present the calculation of the probability of detecting a single infected sample in a pool and in Sec. 5 the calculation of the likelihoods of detecting flawed samples that we then use in the paper. In Sec. 6 we compile some well established formulas with which probabilities can be estimated from frequency of occurrence under the conditions that hold in our cases of interest.

## Acknowledgments

We would like to thank Pablo Aguilar, Alejandro Colaneri and Juliana Sesma for useful discussions and suggestions. This work was partially supported by UBA (UBACyT 20020170100482BA, 20020160100155BA), ANPCyT (PICT 2015-3824, 2015-3583, 2018-02026, 2018-02842), FAPESP (grants 2013/07375-0, 2016/01860-1, and 2018/24293-0) and CNPq (grants 302538/2019-4 and 302682/2019-8).

## References

1. Weissleder R, Lee H, Ko J, Pittet MJ. COVID-19 diagnostics in context. Science Translational Medicine. 2020;12(546). doi:10.1126/scitranslmed.abc1931.

2. Bilder CR. In: Group Testing for Identification. American Cancer Society; 2019. p. 1–11. Available from: https://onlinelibrary.wiley.com/doi/abs/10.1002/9781118445112.stat08227.

3. Bilder CR. In: Group Testing for Estimation. American Cancer Society; 2019. p. 1–11. Available from: https://onlinelibrary.wiley.com/doi/abs/10.1002/9781118445112.stat08231.

4. Dorfman R. The Detection of Defective Members of Large Populations. Ann Math Statist. 1943;14(4):436–440. doi:10.1214/aoms/1177731363.

5. Sterrett A. On the Detection of Defective Members of Large Populations. The Annals of Mathematical Statistics. 1957;28(4):1033–1036.

6. Sobel M, Groll PA. Group testing to eliminate efficiently all defectives in a binomial sample. The Bell System Technical Journal. 1959;38(5):1179–1252. doi:10.1002/j.1538-7305.1959.tb03914.x.

7. Hwang FK. A Method for Detecting All Defective Members in a Population by Group Testing. Journal of the American Statistical Association. 1972;67(339):605–608.

8. Aldridge M. Rates of Adaptive Group Testing in the Linear Regime. 2019 IEEE International Symposium on Information Theory (ISIT). 2019; p. 236–240.

9. Aldridge M, Johnson O, Scarlett J. Group Testing: An Information Theory Perspective. Foundations and Trends in Communications and Information Theory Series. Now Publishers; 2019. Available from: https://books.google.com.ar/books?id=7s5LzAEACAAJ.

10. Phatarfod RM, Sudbury A. The use of a square array scheme in blood testing. Statistics in Medicine. 1994;13(22):2337–2343. doi:10.1002/sim.4780132205.

11. Kim HY, Hudgens MG, Dreyfuss JM, Westreich DJ, Pilcher CD. Comparison of Group Testing Algorithms for Case Identification in the Presence of Test Error. Biometrics. 2007;63(4):1152–1163. doi:10.1111/j.1541-0420.2007.00817.x.

12. McMahan CS, Tebbs JM, Bilder CR. Two-Dimensional Informative Array Testing. Biometrics. 2012;68(3):793–804. doi:10.1111/j.1541-0420.2011.01726.x.

13. Mézard M, Toninelli C. Group testing with random pools: optimal two-stage algorithms. IEEE Trans Inform Theory. 2011;57(3):1736–1745. doi:10.1109/TIT.2010.2103752.

14. Armendáriz I, Ferrari PA, Fraiman D, Martínez JM, Dawson SP. Group testing with nested pools; 2020.

15. Hanel R, Thurner S. Boosting test-efficiency by pooled testing strategies for SARS-CoV-2; 2020.

16. Mentus C, Romeo M, DiPaola C. Analysis and Applications of Non-Adaptive and Adaptive Group Testing Methods for COVID-19. medRxiv. 2020;doi:10.1101/2020.04.05.20050245.

17. Sinnott-Armstrong N, Klein D, Hickey B. Evaluation of Group Testing for SARS-CoV-2 RNA. medRxiv. 2020;doi:10.1101/2020.03.27.20043968.

18. Mutesa L, Ndishimye P, Butera Y, Souopgui J, Uwineza A, Rutayisire R, et al. A pooled testing strategy for identifying SARS-CoV-2 at low prevalence. Nature. 2020;doi:10.1038/s41586-020-2885-5.

19. Mallapaty S. The mathematical strategy that could transform coronavirus testing. Nature. 2020;583:504–505. doi:10.1038/d41586-020-02053-6.

20. Ben-Ami R, Klochendler A, Seidel M, Sido T, Gurel-Gurevich O, Yassour M, et al. Large-scale implementation of pooled RNA extraction and RT-PCR for SARS-CoV-2 detection. Clinical Microbiology and Infection. 2020;26(9):1248–1253. doi:https://doi.org/10.1016/j.cmi.2020.06.009.

21. Shental N, Levy S, Skorniakov S, Wuvshet V, Shemer-Avni Y, Porgador A, et al. Efficient high throughput SARS-CoV-2 testing to detect asymptomatic carriers. medRxiv. 2020;doi:10.1101/2020.04.14.20064618.

22. Barak N, Ben-Ami R, Sido T, Perri A, Shtoyer A, Rivkin M, et al. Lessons from applied large-scale pooling of 133,816 SARS-CoV-2 RT-PCR tests. medRxiv. 2020;doi:10.1101/2020.10.16.20213405.

23. Suo T, Liu X, Feng J, Guo M, Hu W, Guo D, et al. ddPCR: a more accurate tool for SARS-CoV-2 detection in low viral load specimens. Emerging Microbes & Infections. 2020;9(1):1259–1268. doi:10.1080/22221751.2020.1772678.

24. Pilcher CD, Westreich D, Hudgens MG. Group Testing for Severe Acute Respiratory Syndrome– Coronavirus 2 to Enable Rapid Scale-up of Testing and Real-Time Surveillance of Incidence. The Journal of Infectious Diseases. 2020;222(6):903–909. doi:10.1093/infdis/jiaa378.

25. Liu X, Feng J, Zhang Q, Guo D, Zhang L, Suo T, et al. Analytical comparisons of SARS-COV-2 detection by qRT-PCR and ddPCR with multiple primer/probe sets. Emerging Microbes & Infections. 2020;9(1):1175–1179. doi:10.1080/22221751.2020.1772679.

26. Alteri C, Cento V, Antonello M, Colagrossi L, Merli M, Ughi N, et al. Detection and quantification of SARS-CoV-2 by droplet digital PCR in real-time PCR negative nasopharyngeal swabs from suspected COVID-19 patients. PLOS ONE. 2020;15(9):1–10. doi:10.1371/journal.pone.0236311.

27. Suo T, Liu X, Guo M, Feng J, Hu W, Yang Y, et al. ddPCR: a more sensitive and accurate tool for SARS-CoV-2 detection in low viral load specimens. medRxiv. 2020;doi:10.1101/2020.02.29.20029439.

28. Schmid-Burgk JL, Schmithausen RM, Li D, Hollstein R, Ben-Shmuel A, Israeli O, et al. LAMP-Seq: Population-Scale COVID-19 Diagnostics Using Combinatorial Barcoding. bioRxiv. 2020;doi:10.1101/2020.04.06.025635.

29. Vandenberg O, Martiny D, Rochas O, van Belkum A, Kozlakidis Z. Considerations for diagnostic COVID-19 tests. Nature Reviews Microbiology. 2020;doi:10.1038/s41579-020-00461-z.

30. Yelin I, Aharony N, Shaer-Tamar E, Argoetti A, Messer E, Berenbaum D, et al. Evaluation of COVID-19 RT-qPCR test in multi-sample pools. medRxiv. 2020;doi:10.1101/2020.03.26.20039438.

31. Falzone L, Musso N, Gattuso G, Bongiorno D, Palermo CI, Scalia G, et al. Sensitivity assessment of droplet digital PCR for SARS-CoV-2 detection. International Journal of Molecular Medicine. 2020;46(3):957–964. doi:10.3892/ijmm.2020.4673.

32. Hindson BJ, Ness KD, Masquelier DA, Belgrader P, Heredia NJ, Makarewicz AJ, et al. High-Throughput Droplet Digital PCR System for Absolute Quantitation of DNA Copy Number. Analytical Chemistry. 2011;83(22):8604–8610. doi:10.1021/ac202028g.

33. Hindson CM, Chevillet JR, Briggs HA, Gallichotte EN, Ruf IK, Hindson BJ, et al. Absolute quantification by droplet digital PCR versus analog real-time PCR. Nature Methods. 2013;10(10):1003–1005. doi:10.1038/nmeth.2633.

34. Yu F, Yan L, Wang N, Yang S, Wang L, Tang Y, et al. Quantitative Detection and Viral Load Analysis of SARS-CoV-2 in Infected Patients. Clinical Infectious Diseases. 2020;71(15):793–798. doi:10.1093/cid/ciaa345.

35. Zheng S, Fan J, Yu F, Feng B, Lou B, Zou Q, et al. Viral load dynamics and disease severity in patients infected with SARS-CoV-2 in Zhejiang province, China, January-March 2020: retrospective cohort study. BMJ. 2020;369. doi:10.1136/bmj.m1443.

36. Pan Y, Zhang D, Yang P, Poon LLM, Wang Q. Viral load of SARS-CoV-2 in clinical samples. The Lancet Infectious Diseases. 2020;20(4):411–412. doi:https://doi.org/10.1016/S1473-3099(20)30113-4.

