## Supplementary Calculations for "Nested pool testing strategy for the reliable identification of individuals infected with SARS-CoV-2"

#### 1 Nested pooling strategy

The pooling strategy analyzed in the paper is characterized by a number of stages, $k+1$, and a sequence of pool sizes, $m = (m_1, ..., m_k)$, where $m_1 > \cdots > m_k > 1$ and $m_j$ is a multiple of $m_{j+1}$ for all $j$. At the last $(k+1)$ stage the samples are tested individually, *i.e.*, $m_{k+1} = 1$. Given the infection probability, $p$, the optimal strategy is characterized by values, $k$ and $m$, that minimize the cost: the expected number of tests per individual. We present in what follows a brief description of the strategy and the main results obtained in [1] that are used in the paper. An interface to decide the best nested strategy given a set of constraints will be available at https://wp.df.uba.ar/pooling/ for those interested in trying our approach.

##### 1.1 Presentation of the strategy

Let us assume that $N$ individuals have to be tested and that $p$ is the probability that anyone of them is infected. At the first stage we subdivide the $N$ samples into disjoint groups of size, $m_1$, and combine the $m_1$ samples of each group into a single pool that is tested. Once the first-stage tests are performed, we subdivide the samples of each of the pools that detected the presence of at least one infected sample (*i.e.*, that tested positive) into $m_1/m_2$ pools of $m_2$ samples each. We perform the test on these $m_2$ sample pools and proceed as before: those that test positive are subdivided in $m_2/m_3$ pools of $m_3$ samples each. The procedure is iterated until the $(k+1)$-th stage is reached at which all the individual samples of the pools that tested positive at the $k-$th stage are tested. In order to understand the optimization of the procedure it is best to introduce a labeling of the pools at all stages from the very beginning, as if none of them ever tested negative. To this end, let us call, $W_{i_1}$, the set of individuals whose samples are contained in the $i_1-$th pool of the first stage. We subdivide this set into $m_1/m_2$ subsets with $m_2$ individuals each. Let us call $W_{i_1 i_2}$ the sets of individuals whose samples, at the second stage, are in the $i_2-$th subpool of the first stage $i_1-$th pool. We repeat this labeling at all stages, so that at the $k-$th one we have subsets $W_{i_1 \ldots i_k}$. For the sake of simplicity, we will also refer to the pools of samples with these same names, $W_{i_1 \ldots i_k}$. To each of these subsets we assign a variable, $\phi$, such that $\phi = 0$ if there is at least one infected individual in the subset and $\phi = 1$ otherwise. We introduce the notation $\phi_{i_1} = \phi(W_{i_1})$, $\phi_{i_1 i_2} = \phi(W_{i_1 i_2})$, $\ldots$, $\phi_{i_1 \ldots i_k} = \phi(W_{i_1 \ldots i_k})$. We illustrate this labeling in Fig. 1 where we depict one first stage pool, $W_0$, and its subdivision for an example with 4 stages ($k = 3$) that starts with $m_1 = 27$ and such that each (positive) pool is subdivided into three new pools at each successive stage.

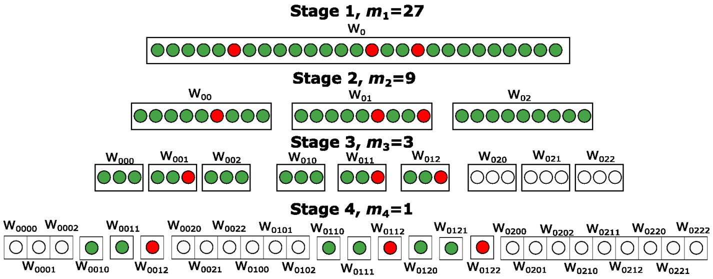


**Fig 1.** Schematic depiction of the nested pool strategy with $k = 3$ and $m = (27, 9, 3)$ applied to an initial pool (rectangle) with samples (circles) coming from the 27 individuals in the set, $W_{i_1}$ with $i_1 = 0$, three of which are infected (red circles in the figure). Each pool of each stage is subdivided into 3 pools at the following stage. The figure illustrates the labels that we assign to the different pools at the various stages and how this can be done *a priori*. In any practical implementation of the strategy, only the pools with infected samples pass to the following stage. In the example of the figure, the test performed on $W_0$ is positive (because there are samples of infected individuals in $W_0$). The subpools, $W_{00}$, $W_{01}$ and $W_{02}$, of $W_0$ are then tested at the second stage, the first two of which turn out to be positive because they contain samples from infected individuals. All the sub-pools of $W_{00}$, $W_{01}$ and $W_{02}$, are depicted in the third-stage row of the figure, but only those that are tested at this stage contain circles that are colored in red or green if infected or not, respectively. The grey circles correspond to samples identified as not infected at previous stages. Thus, only $W_{000}$, $W_{001}$, $W_{002}$, $W_{010}$, $W_{011}$ and $W_{012}$ are tested at the third stage, three of which turn out to be positive so that 9 samples are tested individually at the last ($4^{th}$) stage. The total number of tests that are performed in this case is 19, which is smaller than 27, and the cost is 0.7. As illustrated in Fig. 1 of the paper, this situation occurs in less than 5% of the cases for which the strategy with $k = 3$ and $m = (27, 9, 3)$ is optimal.

#### 1.2 Number of tests

Using the labeling illustrated in Fig. 1 and the definition of $\phi$ we can write the number of tests, $T_k^{(j)}(m, p)$, that are needed to perform at each stage, $2 \leq j \leq k + 1$, of the $(k + 1)$-stage strategy with pool sizes, $m = (m_1, ..., m_k)$, to identify the infected individuals in each first-stage set, $W_{i_1}$. This number is equal to $m_{j-1}/m_j$ times the number of pools that test positive at the $j - 1$ stage:

$$T_k^{(j)}(m, p) = \frac{m_{j-1}}{m_j} \sum_{i_2=0}^{\frac{m_1}{m_2}-1} \sum_{i_3=0}^{\frac{m_2}{m_3}-1} \cdots \sum_{i_{j-1}=0}^{\frac{m_{j-2}}{m_{j-1}}-1} \left(1 - \phi_{i_1 i_2 \ldots i_{j-1}}\right), \qquad 2 \leq j \leq k + 1, \tag{1}$$

where we have introduced $m_{k+1} = 1$. The only non-zero terms in Eq. (1) correspond to the $m_{j-1}$-sample pools with $\phi_{i_1 i_2 \ldots i_{j-1}} = 0$ (*i.e.*, with infected samples). The probability that $\phi_{i_1 i_2 \ldots i_{j-1}} = 0$ is $(1-(1-p)^{m_{j-1}})$ so that the expected value of $T_k^{(j)}(m,p)$ is:

$$ET_k^{(j)}(m,p) = \frac{m_1}{m_j}\big(1-(1-p)^{m_{j-1}}\big), \qquad 2 \le j \le k+1. \tag{2}$$

Taking into account that all pools are tested at the first stage, the expected value of the total number of tests that is needed to identify the infected individuals in any first-stage pool with the $m = (m_1, ..., m_k)$ strategy is then given by:

$$ET_k = 1 + \sum_{j=2}^{k} \frac{m_1}{m_j}\big(1-(1-p)^{m_{j-1}}\big) + m_1\left(1-(1-p)^{m_k}\right). \tag{3}$$

The variance of the number of tests can also be computed analytically [1]. We only quote here the result for the special case of the strategies in which the ratio of consecutive pool sizes is constant and equal to $m_k$. As described later, a strategy of this type (with $m_k = 3$) is the optimal for most values of the infection probability, $p$. The variance for the strategies with $m = (\mu^k, \mu^{k-1}, \ldots, \mu)$, $\mu > 1$, can be written as [1]:

$$\begin{aligned}\mathrm{Var}\big(T_k\big) &= \mu^2\Big\{\sum_{i=1}^{k}\mu^{i-1}\big(1-q^{m_i}\big)\Big[q^{m_i} + 2\sum_{j=1}^{i-1} q^{m_j}\Big]\Big\} &(4)\\ &= \mu^2\Big\{\sum_{i=1}^{k}\mu^{i-1}\Big(1-q^{\mu^{k-i+1}}\Big)\Big[q^{\mu^{k-i+1}} + 2\sum_{j=1}^{i-1} q^{\mu^{k-j+1}}\Big]\Big\}, &(5)\end{aligned}$$

where $q = 1-p$. This variance gives a standard deviation for the number of tests per individual:

$$\sigma(m,p) = \frac{1}{\mu^k}\sqrt{\mathrm{Var}\big(T_k(m,p)\big)}. \tag{6}$$

Given a nested strategy, the probability of having to perform a certain number of tests can also be computed analytically as a function of the infection probability, $p$, using combinatorics. The problem becomes increasingly complicated as the size of the initial pool gets larger and the number of possible outcomes also increases. We hereby describe how to proceed in the case of the strategy illustrated in Fig. 1, which has $k=3$ and $m = (27, 9, 3)$. Given an initial pool, the total number tests, $T$, that will have to be performed under such strategy can take on ten possible values:
$\{1, 7+3\times 1, 7+3\times 2, \ldots, 7+3\times 9\}$. The procedure consists in counting in how many ways one can end up performing a number of tests, $T$, equal to one of these ten values, given a number of infected samples that can go from 0 through 27, pondering the various cases by their probability of occurrence. We limit the description to computing the probabilities of occurrence of the first few possibilities, but the procedure can be extended to all. Let us then call *minimal subpools* the sample pools associated to the nine sets, $W_{i_1 i_2 i_3}$, $0 \le i_1, i_2, i_3 \le 2$ (third row in the scheme of Fig. 1) and *maximal subpools* those associated to the three sets $W_{i_1 i_2}$, $0 \le i_1 i_2 \le 2$ (second row in Fig. 1).

The first case, $T = 1$, occurs when there are no infected samples in the initial pool. This occurs with probability:

$$P(T=1) = (1-p)^{27}. \tag{7}$$

The second case, $T = 10$, occurs when all the infected samples of the pool belong to the same minimal subpool. A minimal subpool can have 1, 2 or 3 infected samples. To compute the probability we first fix the number of infected samples and then count in how many ways we can distribute them so that they all belong to the same minimal subpool. Finally we add over the situations with 1, 2 or 3 infected samples with their corresponding probability of occurrence. We obtain:

$$P(T=10)=\sum_{\ell=1}^{3}\binom{3}{1}\binom{3}{1}G^a_\ell p^\ell(1-p)^{27-\ell} \tag{8}$$

In this equation, the term, $\binom{3}{1}\binom{3}{1}$, corresponds to the number of ways in which a minimal subpool can be selected. The term, $G^a_\ell := \binom{3}{\ell}$, is the number of different configurations with $\ell$ infected samples in the minimal subpool selected. The factor, $p^\ell(1-p)^{27-\ell}$, is the probability of having $\ell$ infected samples. The third case, $T = 13$, occurs if all the infected samples are in exactly two minimal subpools that belong to the same maximal subpool. In this case,

$$P(T=13)=\sum_{\ell=2}^{6}\binom{3}{1}\binom{3}{2}G^b_\ell p^\ell(1-p)^{27-\ell}. \tag{9}$$

The term, $\binom{3}{1}\binom{3}{2}$, represents the number of ways in which two minimal subpools that belong to the same maximal subpool can be selected. The term, $G^b_\ell := \binom{6}{\ell} - 2\binom{3}{\ell}$, is the number of different configurations with $\ell$ positve infected samples in the subpool selected, considering $\binom{n}{\ell} = 0$ if $k > \ell$. $T$ takes on the value 16 if all the infected samples are either in exactly three minimal subpools that belong to the same maximal subpool, or they are in exactly two minimal subpools that belong to two different maximal subpools. In this case:

$$P(T=16)=\sum_{\ell=2}^{6}\binom{3}{2}\binom{3}{1}\binom{3}{1}G^b_\ell p^\ell(1-p)^{27-\ell}+\sum_{\ell=3}^{9}\binom{3}{1}\binom{3}{3}G^c_\ell p^\ell(1-p)^{27-\ell}, \tag{10}$$

with $G^c_\ell = \binom{9}{\ell} - (\binom{3}{2}\binom{6}{\ell} - 3\binom{3}{\ell})$. Eqs (7)–Eqs (10) were used to compute the analytic results depicted in Fig. 2 of the paper.

The probability of the number of infected samples in a pool of the first ($N_i^{(1)}$), second ($N_i^{(2)}$) or third stages ($N_i^{(3)}$) can be computed similarly. In particular, for a first stage pool (with $m_1 = 27$) it is:

$$P(N_i^{(1)}=\ell)=\binom{27}{\ell}p^\ell(1-p)^{27-\ell}, \quad \ell=\{0,1,\ldots,27\}. \tag{11}$$

The computation for $N_i^{(2)}$ and $N_i^{(3)}$ is cumbersome. In order to simplify it, we compute the probabilities subject to the condition $0 < N_i^{(1)} \le 4$ (instead of requesting only that $0 < N_i^{(1)}$) which is reasonable to compare it with the results of Fig. 3 of the paper. The computation is straightforward. We obtain:

$$\begin{aligned}P(N_i^{(2)}=0|0<N_i^{(1)}\le 4)&=\frac{1}{P(0<N_i^{(1)}\le 4)}\times[\frac{2}{3}\binom{27}{1}p^1(1-p)^{26}+\\ &\left(\frac{2}{3}\binom{3}{1}\binom{9}{2}+\frac{1}{3}\binom{3}{2}\binom{9}{1}\binom{9}{1}\right)p^2(1-p)^{25}+\left(\frac{2}{3}\binom{3}{1}\binom{9}{3}+\frac{1}{3}2\binom{3}{2}\binom{9}{2}\binom{9}{1}\right)p^3(1-p)^{24}\\ &+\left(\frac{2}{3}\binom{3}{1}\binom{9}{4}+\frac{1}{3}\binom{3}{2}\left(2\binom{9}{1}\binom{9}{3}+\binom{9}{2}\binom{9}{2}\right)\right)p^4(1-p)^{23}],\end{aligned} \tag{12}$$

$$P(N_i^{(2)}=1|0<N_i^{(1)}\le 4)=\frac{1}{P(0<N_i^{(1)}\le 4)}\times\big[\frac{1}{3}\binom{27}{1}p^1(1-p)^{26}+$$
$$\frac{2}{3}\binom{3}{2}\binom{9}{1}\binom{9}{1}p^2(1-p)^{25}+\left(\binom{3}{1}\binom{3}{1}\binom{3}{1}+\frac{1}{3}2\binom{3}{2}\binom{9}{2}\binom{9}{1}\right)p^3(1-p)^{24}$$
$$+\left(\frac{1}{3}\binom{3}{2}\left(2\binom{9}{1}\binom{9}{3}+\frac{2}{3}\binom{3}{1}\binom{9}{2}\binom{9}{1}\binom{9}{1}\right)\right)p^4(1-p)^{23}\big],\tag{13}$$

$$P(N_i^{(2)}=2|0<N_i^{(1)}\le 4)=\frac{1}{P(0<N_i^{(1)}\le 4)}\times\big[\frac{1}{3}\binom{3}{1}\binom{9}{2}p^2(1-p)^{25}+$$
$$\frac{1}{3}\binom{3}{1}\binom{9}{2}\binom{9}{1}p^3(1-p)^{24}+\left(\frac{1}{3}\binom{3}{1}\binom{9}{2}\binom{9}{1}\binom{9}{1}+\frac{2}{3}\binom{3}{2}\binom{9}{2}\binom{9}{2}\right)p^4(1-p)^{23}\big]\tag{14}$$

$$P(N_i^{(3)}=0|0<N_i^{(1)}\le 4)=\frac{1}{P(0<N_i^{(1)}\le 4)}\times\big[\frac{2}{3}\binom{27}{1}p(1-p)^{26}+$$
$$\left(\frac{2}{3}\binom{3}{1}\binom{3}{2}+\frac{2}{3}\binom{3}{2}\binom{9}{1}\binom{9}{1}+\frac{1}{3}\binom{3}{2}\binom{3}{1}\binom{3}{1}\right)p^2(1-p)^{25}+$$
$$\left(\frac{2}{3}\binom{9}{1}\binom{9}{1}\binom{9}{1}+\frac{1}{2}2\binom{9}{2}\binom{3}{2}\binom{3}{1}\right)p^3(1-p)^{24}+$$
$$\left(\frac{1}{3}\binom{9}{2}\binom{3}{2}\binom{3}{2}+\frac{2}{3}\binom{9}{4}\binom{3}{1}\binom{3}{1}\binom{3}{1}\binom{3}{1}\right)p^4(1-p)^{23}\big]\tag{15}$$

$$P(N_i^{(3)}=1|0<N_i^{(1)}\le 4)=\frac{1}{P(0<N_i^{(1)}\le 4)}\times\big[\frac{1}{3}\binom{27}{1}p(1-p)^{26}+$$
$$\frac{2}{9}\binom{9}{2}\binom{3}{1}\binom{3}{1}p^2(1-p)^{25}+(\frac{1}{9}2\binom{9}{2}\binom{3}{2}\binom{3}{1}+\frac{3}{9}\binom{9}{3}\binom{3}{1}\binom{3}{1}\binom{3}{1})p^3(1-p)^{24}+$$
$$\frac{1}{9}\binom{9}{2}2\binom{3}{3}\binom{3}{1})p^4(1-p)^{23}\big]\tag{16}$$

$$P(N_i^{(3)}=2|0<N_i^{(1)}\le 4)=\frac{1}{P(0<N_i^{(1)}\le 4)}\times\big[\frac{1}{3}\binom{3}{1}\binom{3}{1}\binom{3}{2})p^2(1-p)^{25}+$$
$$(\frac{1}{3}2\binom{3}{1}\binom{3}{1}\binom{3}{2}\binom{3}{1}+\frac{1}{6}\binom{3}{2}\binom{3}{1}\binom{3}{1}\binom{3}{1}\binom{3}{2})p^3(1-p)^{24}+$$
$$(\frac{1}{3}2\binom{3}{1}\binom{3}{2}\binom{3}{3}\binom{3}{1}+\frac{2}{6}\binom{3}{2}\binom{3}{1}\binom{3}{1}\binom{3}{2}\binom{3}{2}+\frac{1}{9}\binom{3}{3}\binom{3}{1}\binom{3}{2}\binom{9}{1}\binom{9}{1})p^4(1-p)^{23}\big]\tag{17}$$

##### 1.3 Cost and optimization

The aim of pooling is to reduce the number of tests as much as possible. We then define the *cost* as the expected number of tests per individual:

$$D_k(m,p)\equiv\frac{1}{m_1}ET_k,\tag{18}$$

which, given Eq. (3), is equal to:

$$D_k(m,p)=\frac{1}{m_1}+\sum_{j=2}^{k}\frac{1}{m_j}\left(1-(1-p)^{m_{j-1}}\right)+1-(1-p)^{m_k}.\tag{19}$$

Some properties are derived from the linearization in $p$ of Eq. (19) which is given by:

$$\overline{D}_k(m_1, \ldots, m_k, p) = \frac{1}{m_1} + m_k p + p \sum_{j=2}^{k} \frac{m_{j-1}}{m_j}. \tag{20}$$

This expression is similar to the expected number of tests per individual for realizations with at most one infected sample per initial pool if we replace $p$ by the infection prevalence, $\overline{p}$. We proved [1] that:

$$D_k(m_1, \ldots, m_k, p) \leq \overline{D}_k(m_1, \ldots, m_k, p). \tag{21}$$

This implies that we can always expect our proposed strategy to produce, on average, a larger reduction in the number of tests per individual than the one predicted by Eq. (20).

The optimization problem consists in finding the value, $k$, and the sequence of pool sizes, $m$, that minimize (19) for a given $p$. We proved in [1] that, for $p \geq 1 - 3^{-1/3}$ it is best not to pool and that, for $p \in (2^{-51}, 1 - 3^{-1/3})$, the optimal nested strategy is of the form $(3^k, \ldots, 3)$ for most values of $p$ interspersed with small $p$ intervals over which it is $(3^{k-1}4, 3^{k-1}, \ldots, 3)$. In [1] we gave a precise description of the optimal values of $k$ as functions of $p$ and of the $p$-intervals over which one or the other scheme is best. We show the cost of the optimal strategy as a function of $p$ in Fig. 2 (a). We conjecture it holds true for all $p \in (0, 1)$.

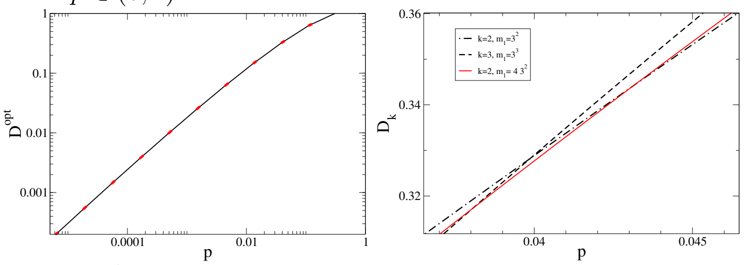


**Fig 2.** (a) Cost, $D^{opt}$, of the optimal nested strategy as a function of $p$ in log-log scale. Red regions correspond to strategies with $m_1 = 4 \times 3^{k-1}$, the rest to $m_1 = 3^k$. We observe that, for every order of magnitude in $p$, the most likely best nested strategy is the one with $m = (3^k, ..., 3)$. (b) Cost of the strategies with $m = (3^2, 3)$ (black dashed-dotted curve), $m = (4 \cdot 3^2, 3)$ (red solid curve) and $m = (3^3, 3^2, 3)$ (black dashed curve). We observe that for the largest (smallest) probabilities displayed in the figure, the first (third) one is best. In between there is a small interval over which the best is the one with $m_1 = 4 \cdot 3^2$.

The costs of the strategies with $m = (3^3, 3^2, 3)$ (black dashed curve), $m = (4 \cdot 3^2, 3)$ (red solid curve) and $m = (3^2, 3)$ (black dashed-dotted curve) are shown in Fig. 2 (b). These strategies are optimal for $0.0146 \lesssim p \lesssim 0.0380$; $0.0380 \lesssim p \lesssim 0.0431$ and $0.0431 \lesssim p \lesssim 0.1098$, respectively [1]. Not only the interval of optimality for the strategy with $m_1 = 4 \cdot 3^2$ is very small compared to those of the other two (Fig. 2 (a)), but also the difference between the three costs is very small ($< 0.4\%$) over this interval. A similar behavior is observed for all $k$.

Restricting the optimization to a search over the strategies with $m = (\mu^k, \ldots, \mu)$, the optimal $k$ as a function of $p$ is given by [1]:

$$k_\mu(p) \equiv \left\lfloor \log_\mu\left(\frac{1}{\log_\mu(1/(1-p))}\right)\right\rfloor = \left\lfloor \log\left(\frac{\log(\mu)}{\log(1/(1-p))}\right)/\log(\mu)\right\rfloor, \tag{22}$$

where $\lfloor \zeta \rfloor$ is the largest integer, $n_\zeta$, such that $n_\zeta \leq \zeta$. As mentioned in the paper, we proved in [1] that the cost of the strategy with $m = (3^{k_3(p)}, ..., 3)$ and $k_3(p)$ given by

Eq. (22) and $\mu = 3$ is not only the optimal one for most values of $p$ but it is also of the same order of magnitude, $O\big(p\log(1/p)\big)$, as the cost of the optimal one for the rest of the values of $p$ (as illustrated in Fig. 2. That is why in the paper we mostly focus on the strategies of the form $m = (3^k, ..., 3)$.

###### 1.3.1 Algorithm to search for the optimal strategy when there are constraints

Although we have a prescription of the optimal nested strategy for every infection probability, $p > 2^{-51}$, sometimes there are constraints that prevent the optimal from being used. In such a case, an algorithmic approach can be used to search for the optimal strategy. The idea is to build a "tree" of sequences as described in what follows. To simplify the description we here characterize the (candidate) nested strategies by the sequence of numbers $m = (m_1, \ldots, m_k, m_{k+1})$ with $m_{k+1} = 1$, *i.e.*, with an additional 1 at the end that represents the stage of individual sample testing. The tree is then constructed as follows:

1. The root of the tree is the sequence with a single element, $(m_1)$.
2. If the sequence $(m_1, \ldots, m_\ell)$ is a node of the tree and $m_\ell \neq 1$, there are as many branches emerging from this node as the number of proper submultiples of $m_\ell$ plus one. Here by proper we mean the natural numbers, $m_{\ell,1}, \ldots, m_{\ell,q}$, such that the ratios $m_\ell/m_{\ell,s}$, $1 \leq s \leq q$, are integers different from one. The nodes that branch out from $(m_1, \ldots, m_\ell)$ are: $(m_1, \ldots, m_\ell, m_{\ell,1}), \ldots (m_1, \ldots, m_\ell, m_{\ell,q})$, and $(m_1, \ldots, m_\ell, 1)$.
3. If the sequence $(m_1, \ldots, m_\ell)$ is a node of the tree and $m_\ell = 1$ there are no branches emerging from $(m_1, \ldots, m_\ell)$. This node then corresponds to a fesible nested strategy with $k = \ell - 1$.

As a consequence of this construction the terminal nodes of the tree are all the feasible sequences that start with the number, $m_1$, of the root. The cost, $D_k(m, p)$, of each of these sequences can be computed using Eq. (19). The sequence with the minimal value of $D_k(m, p)$ is the optimal for the given $m_1$ and $p$.

The number of nodes can be reduced by taking into consideration the results of two lemmas that were proved in [1]. Namely, that at an optimal solution with $m = (m_1, ..., m_k)$: *i)* the ratio $m_j/m_{j+1}$ is not smaller than the ratio $m_{j+1}/m_{j+2}$ for all $1 \leq j \leq k-1$; *ii)* $m_j/m_{j+1}$ can only be equal to two for $j = k$. These two lemmas prune the tree and accelerate the search of the optimal strategy for a given $m_1$.

#### 2 Estimating the infection probability, $p$.

The optimal strategy just described depends on the infection probability, $p$, a parameter that might be unknown *a priori*. As shown in the paper, applying a strategy that is optimal for a given $p$ to a situation with smaller infection probability still produces a noticeable reduction in the number of tests per individual that have to be performed to identify the infected samples. The fact that PCR tests can be run in parallel on several (individual or pools of) samples at the same time using multi-well plates opens the possibility of applying a sub-optimal strategy and using the first round of tests to obtain an estimate of $p$. In this way, one option is to start with the strategy that is optimal for the largest $p$ for which the pooling of samples is advisable ($p \sim 0.3$). The infected samples identified in this way could give an estimate of $p$ with which to choose the strategy to be applied to the following batch of pools. Now, this largest $p$ optimal strategy starts with pools of $m_1 = 3$ samples. Thus, if $p$ is moderately small it is likely that, when running the strategy in parallel on a 96-well plate, no pool will test positive ($\sim 1.4$ pools would test positive on average for $p = 0.005$). In such a case the first (parallel) run would not provide an estimate of $p$. Another possibility is to perform a first (parallel) round solely to estimate $p$. In either case, these runs would allow the identification of some of the non-infected samples (those that happen to be in pools with no infected samples). We describe in what follows how we can proceed if we choose one or the other option.

#### 2.1 Running a set of parallel tests to infer $p$.

Let us assume that we are working with a 96-well plate. Situations with plates of other sizes can be handled analogously.

1. We fill 32 pools with $m_1 = 3$ samples, another 32 pools with $m_1 = 9$ and the last 32 with $m_1 = 27$ samples.

2. We run the test on the 96 pools simultaneously and determine the number, $W_h$, of pools with $m_1 = h$ ($h = 3, 9, 27$) that test positive in this first run.

3. We compute the estimator of the probability, $\hat{P}_h$, that a pool with $m_1 = h$ samples within a set of $N_w = 32$ pools turns out to be positive as:

$$\hat{P}_h = \frac{W_h}{N_w + \frac{h-1}{2h}}. \tag{23}$$

4. We derive an estimator, $\hat{p}$, of the (individual) sample infection probability, $p$, as:

$$\hat{p} = \frac{1}{3}\left(\hat{p}_3 + \hat{p}_9 + \hat{p}_{27}\right), \tag{24}$$

   where

$$\hat{p}_h = 1 - (1 - \hat{P}_h)^{1/h}; \quad h = 3, 9, 27. \tag{25}$$

   The term $\frac{h-1}{2h}$ is added in order to reduce the bias [2]. If any of the values, $\hat{p}_h$, is too small, it is best to follow the procedure described in the next section.

#### 2.2 Dynamic estimation of $p$ as the strategy is applied

In this case we apply the strategy as stated in the manuscript, with some modifications depending on the outcome of the first set of parallel tests. If $p$ is completely unknown but one wants to apply the strategy anyway, it is best to start with $m_1 = 3$. The test is then run simultaneously on 96 pools with 3 samples each. There are four possible types of outcomes and, thus, of subsequent steps, as described in what follows.

I) *If the number of 3-sample pools that test positive in the first run, $N_p^+$, is such that $0 < 3N_p^+ < 96$:*

1. We obtain an estimate, $\hat{p}$, of the individual sample infection probability as $\hat{p} = \hat{p}_h$ using Eqs. (23) and (25) with $N_w = 96$, $h = 3$ and $W_h = N_p^+$. We derive the optimal pool size for the estimated probability, $m_1(\hat{p})$, as:

$$m_1(\hat{p}) = 3^{\hat{k}_3}, \text{with} \quad \hat{k}_3 = \left\lfloor \log_3\left(\frac{1}{\log_3(1/(1-\hat{p}))}\right)\right\rfloor. \tag{26}$$

2. We accommodate, in the 96-well plate, the $3N_p^+$ samples of the $N_p^+$ pools that tested positive in the first stage and $96 - 3N_p^+$ pools with $m_1 \leq m_1(\hat{p})$ samples each. If $m_1(\hat{p})$ is smaller than the maximum number allowed for a reliable detection, then we set $m_1 = m_1(\hat{p})$, if not we choose the largest number of the form $m_1 = 3^k$ that allows the reliable detection of one infected sample in an $m_1$-pool.

3. After the second run of 96 parallel tests is done we know the number, $N_i$, of infected samples that were contained in the first set of 96 3-sample pools. We can then obtain a better estimate of the infection probability as:

$$\hat{p} = \frac{N_i}{96h}, \tag{27}$$

with $h = 3$. Using this estimate we recompute the best pool size, $m_1$, using Eq. (26).

4. For the third run of tests, we use as many wells as necessary to accommodate the $(m_1/3)$-sample pools that come from the $m_1$-sample pools that tested positive in the second run and, if there is room, pools with the newly determined value of $m_1$. This procedure is then continued as long as there are samples.

II) *If the number of 3-sample pools that test positive in the first run,* $N_p^+$*, is such that* $96 \leq 3N_p^+ < 3 \cdot 63 = 189$*:*

1. We obtain an estimate, $\hat{p}$, of the individual sample infection probability as in the previous case, namely using Eqs. (23) and (25) to compute $\hat{p}_h$ with $h = 3$ and then set $\hat{p} = \hat{p}_h$. We then use Eq. (26) to estimate the best pool size, $m_1(\hat{p})$.

2. We test individually 96 of the $3N_p^+$ samples contained in the $N_p^+$ pools that tested positive in the first run. We keep on doing this with the other individual samples of the initial $N_p^+$ pools as long as there are no unused wells in the plate. Once there is room, we pool the new samples in groups of $m_1$ samples each with $m_1$ given by the estimate, $m_1(\hat{p})$, derived in the previous step if this value does not exceed the maximum allowed for a reliable detection. Otherwise, we choose, as before, the largest $m_1 = 3^k$ that satisfies the restriction for a reliable detection.

3. Once all the samples contained in the initial 3-sample pools are identified as infected or not, we know the number, $N_i$, of infected samples in the initial batch with which we can obtain a better estimate of the infection probability using Eq. (27) with $h = 3$ and then compute the best pool size, $m_1$, with Eq. (26).

III) *If the number of 3-sample pools that test positive in the first run,* $N_p^+$*, is such that* $3N_p^+ \geq 3 \cdot 63 = 189$*:*

1. We continue working with individual samples (no pooling) both with those contained in the $N_p^+$ pools that tested positive in the first run and with the untested samples so far. For the latter, each run will give the number of infected individuals, $N_i$, from which the infection probability, $\hat{p}$, can be estimated using Eq. (27) with $h = 1$. As long as the obtained value satisfies $\hat{p} \geq 1 - 1/3^{1/3} \approx 0.307$ we continue without pooling. Otherwise, the optimal $m_1$ can be determined with Eq. (26) and a strategy that starts with this value of $m_1$ (or smaller as discussed before) is applied.

IV)*If none of the 3-sample pools tests positive:*

1. We choose $m_1$ as the the maximum number of the form $m_1 = 3^k$ that allows the reliable detection of one infected sample within an $m_1$ pool.

If an estimate, $\hat{p}$, of the infection probability is available *a priori*, we then start with pools of size $m_1$ as defined in Eq. (26). If only the order of magnitude of $\hat{p}$ is known, we then start with $m_1 = 3^{\hat{k}_3 - 1}$ with $\hat{k}_3$ as in Eq. (26). After the first run of tests is performed on the 96 pools containing $m_1$ samples each, we compute the estimator, $\hat{P}_h$, of the probability that a pool tests positive using Eq. (23) with $N_w = 96$ and $h = m_1$. The individual sample probability is then estimated using Eq. (25) with $h = m_1$. This newly determined estimate, $\hat{p}$, is then used to decide the size of the new pools if there is room to place them in the plate together with those that come from the 96 pools tested in the first run, as explained in II) before. Once the number, $N_i$, of infected samples among the $96m_1$ tested in the first run is known, a new estimate of the infection probability can be obtained using Eq. (27) with $h = m_1$. The infection probability can be re-estimated as the strategy is applied either using Eqs. (23) and (25) or Eq. (27), depending on whether we use the information drawn, respectively, from the first or the last stage of the strategy. Averaging the estimates obtained in various ways could also be a good option.

### 3 Estimating nucleic acid content using ddPCR

#### 3.1 The general case

In ddPCR the volume of each test, *i.e.*, the testing volume, $V_t$, that goes into one well is divided into $M$ sub-volumes, $V_{gt}$, of $\sim 1nl$ each:

$$V_t = MV_{gt}. \tag{28}$$

$M$ varies depending on the equipment. We will use $M = 20,000$, $V_{gt} = 1nl$ and $V_t = 20\mu l$. The testing volume, $V_t$, in turn, comes from a dilution of a volume, $V_s$, that is taken from the sample. Namely, $V_t$ contains the nucleic acid molecules contained in a volume, $V_s$, taken from the original sample and other solutions with the material that is needed for the test to proceed. Given that this procedure does not add any new molecules of the nucleic acids that the test identifies, for the purpose of their quantification we merely think of it as introducing a dilution of factor, $D$, so that:

$$V_t = DV_s. \tag{29}$$

$V_s$, on the other hand, is part of the volume of biological material that is taken from the individual to detect the presence of the nucleic acids of interest. We call, $V_o$, the volume of this biological material of which $V_s$ is a sub-volume with no dilutions involved. We clarify what we mean by this in what follows. The aim of the quantification is to determine the concentration, $c_R$, of the nucleic acids in $V_o$. To this end, we think of $V_s$ as the sum of $M = 20,000$ sub-volumes, $V_{gs}$, each of which, when diluted by the factor $D$, corresponds to one of the sub-volumes of the ddPCR test:

$$V_s = MV_{gs}, \tag{30}$$

$$V_{gt} = DV_{gs}. \tag{31}$$

We assume that each of the sub-volumes, $V_{gs}$, is a sufficiently small fraction of $V_o$ so that the numbers of nucleic acid molecules contained in each of them, $N_{Rg}$, are independent identically distributed (i.i.d.) random variables. The mean of these variables is such that

$$\langle N_{Rg} \rangle = c_R V_{gs} = \frac{c_R}{D} V_{gt}. \tag{32}$$

The first of the two equalities in Eq. (32) reflects what we meant before when we said that there was no dilution involved in the extraction of $V_s$ from $V_o$. Given that the dilution that transforms $V_s$ into $V_t$ does not change the nucleic acid content, these i.i.d. random variables also correspond to the numbers of nucleic acid molecules in each of the tested sub-volumes, $V_{gt}$. The variables, $N_{Rg}$, are described by a Binomial distribution:

$$N_{Rg} \sim B(N_{Ro}, p_{gs}), \tag{33}$$

of parameters

$$N_{Ro} = c_R V_o, \quad p_{gs} = \frac{V_{gs}}{V_o} = \frac{V_{gt}}{D V_o}. \tag{34}$$

As we show later, the Poisson approximation with mean given by Eq. (32) serves very well for our purpose too. ddPCR is an end-point method which determines (ideally) how many of the $M$ $V_{gt} \sim 1nl$ sub-volumes contained at least one nucleic acid molecule. Under the assumption that the $M$ numbers, $N_{Rg}$, are i.i.d. according to Eqs. (33)-(34), the fraction of sub-volumes with at least one nucleic acid molecule is an estimator of the probability, $P_{+g}$, that one such volume contains at least one of these molecules:

$$P_{+g} = 1 - P(N_{Rg} = 0) = 1 - (1 - p_{gs})^{N_{Ro}}. \tag{35}$$

Using the second equation in (34) we can rewrite Eq. (35) as:

$$P_{+g} = 1 - \left(1 - \frac{V_{gs}}{V_o}\right)^{N_{Ro}}, \tag{36}$$

or, equivalently, as:

$$\log\left(1 - \frac{V_{gs}}{V_o}\right) = \frac{1}{N_{Ro}} \log\left(1 - P_{+g}\right). \tag{37}$$

Given Eq. (30) and the fact that $V_s < V_o$ it is $V_{gs}/V_o < 1/M = 5\,10^{-5}$, the l.h.s. of Eq. (37) can be approximated by $-V_{gs}/V_o$. With this approximation and using the two equations in (34), Eq. (37) can be rewritten as:

$$c_R \frac{V_{gt}}{D} = -\log\left(1 - P_{+g}\right). \tag{38}$$

This equation is also obtained if the variables, $N_{Rg}$, are assumed to be Poisson distributed with mean given by Eq. (32) in which case $1 - P_{+g} = \exp(-c_R V_{gt}/D)$.

Let us assume that the test determined that $M_+$ of the $M$ sub-volumes contained molecules of the nucleic acid of interest. As explained in Sec. 3.3, if $MP_{+g} > 5$ and $M(1 - P_{+g}) > 5$, the fraction:

$$\hat{P}_{+g} = \frac{M_+}{M}, \tag{39}$$

is an estimator of $P_{+g}$ such that:

$$P_{+g} \in \left[\hat{P}_{+g} - 1.96\sqrt{\frac{\hat{P}_{+g}\left(1 - \hat{P}_{+g}\right)}{M}}, \hat{P}_{+g} + 1.96\sqrt{\frac{\hat{P}_{+g}\left(1 - \hat{P}_{+g}\right)}{M}}\right], \tag{40}$$

with 95% confidence. If not, other intervals for $P_{+g}$ can be estimated as explained in Sec. 3.3. The "large sample" limit ($MP_{+g} > 5$ and $M(1 - P_{+g}) > 5$) guarantees that the borders of the interval are bounded away from 0 and 1. Another important property of this limit is that $\hat{P}_{+g}$ is normally distributed around $P_{+g}$. We can then replace $P_{+g}$ in Eq. (38) by its estimator, $\hat{P}_{+g}$, [3] and obtain an estimate, $\hat{c}_R$, of the nucleic acid concentration in the original sample, $c_R$, as:

$$\hat{c}_R \frac{V_{gt}}{D} = -\log\left(1 - \hat{P}_{+g}\right). \tag{41}$$

This is the usual formula with which the content of nucleic acids is quantified in ddPCR experiments (see *e.g.*, [3]). In view of Eq. (40), the range of possible values, $c_R$, for each value of $\hat{P}_{+g}$ is then given by:

$$-\log\left(1 - \hat{P}_{+g} + 1.96\sqrt{\frac{\hat{P}_{+g}\left(1-\hat{P}_{+g}\right)}{M}}\right) \leq c_R \frac{V_{gt}}{D} \leq -\log\left(1 - \hat{P}_{+g} - 1.96\sqrt{\frac{\hat{P}_{+g}\left(1-\hat{P}_{+g}\right)}{M}}\right). \tag{42}$$

Fig. 3 illustrates how the range of $c_R$ values given by Eq. (42) and the correspondig relative uncertainty behave as functions of $\hat{P}_{+g} = M_+/M$ for values of $\hat{P}_{+g}$ for which the large sample limit holds as described in what follows.

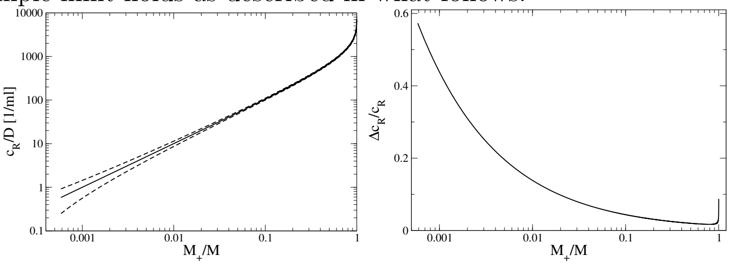


**Fig 3.** (a) Predictor, $\hat{c}_R/D$, of the nucleic acid concentration in the testing well derived from Eq. (41) (solid line) and upper and lower limits of the values that the same concentration, $c_R/D$, can on with a 95% confidence level derived from Eq. (42) (dashed lines) as functions of the experimentally determined, $\hat{P}_{+g} = M_+/M$, for $\hat{P}_{+g}$ within the range defined in Eq. (43). For all computations we used $M = 20,000$ and $V_{gt} = 10^{-3} ml$. (b) Relative uncertainty of the predicted nucleic acid concentration, $c_R/D$, (*i.e.* one half of the difference between the upper and lower limits plotted with dashed lines in (a) divided by the predicted value, $\hat{c}_R/D$, of Eq. (41))as a function of $\hat{P}_{+g} = M_+/M$.

Given Eq. (40), the conditions for the large sample limit to hold, $MP_{+g} > 5$ and $M(1 - P_{+g}) > 5$, are satisfied if $\left(\hat{P}_{+g} - 1.96\sqrt{\hat{P}_{+g}(1-\hat{P}_{+g})/M}\right) > 5/M$ and $\left(1 - \hat{P}_{+g} - 1.96\sqrt{\hat{P}_{+g}(1-\hat{P}_{+g})/M}\right) > 5/M$ from which we obtain:

$$5.85\,10^{-4} < \hat{P}_{+g} < 0.999415. \tag{43}$$

This implies that the large sample limit can be used if the number of sub-volumes that test positive satisfy $11 < M_+ < 19989$. In terms of the estimated concentration, Eq. (43) can be rewritten as:

$$\frac{5.85\,10^{-4}}{V_{gt}} < \frac{\hat{c}_R}{D} < \frac{7.44}{V_{gt}}, \tag{44}$$

or, equivalently, that

$$\frac{11.7}{V_t} < \frac{\hat{c}_R}{D} < \frac{1.49\,10^5}{V_t}, \tag{45}$$

$$\frac{585}{ml} < \frac{\hat{c}_R}{D} < \frac{7.4\,10^6}{ml}, \tag{46}$$

where we have used that the testing sample, $V_t = 20\mu l$, is such that $V_t = 20,000 V_{gt}$. As discussed in the paper, these ranges allow the quantification of viral RNA in most of the samples taken from people infected with SARS-CoV-2, with the exception of those with the very highest viral loads [4–7]. As illustrated in Fig. 3, the uncertainty in the determination of the concentration, on the other hand, remains at manageable levels throughout the range for which the large sample limit holds. Even though at very low values the uncertainty is $\sim 60\%$, which implies that only its order of magnitude can be determined when $c_R/D \sim \mathcal{O}(10^{-1}/ml)$, at $M_+/M \sim 0.005$ the uncertainty is already of the order of 20%. For larger, $M_+/M$, the uncertainty goes down and never exceeds 10%.

#### 3.2 The case of pooled samples.

Let us apply now the analysis just described to the case of pools of samples. To this end, let us denote by *original sample* the sample that is taken from an individual before it is diluted and/or mixed with any other sample to proceed with the test. Let us identify each of the original samples that go in the pool of interest with the subscript, $i$, $1 \le i \le m_j$. Each of these original samples has a volume, $V_{o;i}$ and a certain concentration, $\tilde{c}_{R;i}$, of the RNA that is to be detected by the test ($\tilde{c}_{R;i} = 0$ is an option). Let us assume that, for the test, a volume, $V_{s;i}^{(j)}$, is drawn from $V_{o;i}$ for each $1 \le i \le m_j$. We assume that the volume extraction is done in such a way that guarantees a small variability of $V_{s;i}^{(j)}$ among samples so that $V_{s;i}^{(j)}$ has a fixed value, $V_s^{(j)} = V_s/m_j$, for all values of $i$. The volumes, $V_{s;i}^{(j)}$, $1 \le i \le m_j$, are at some point combined into one volume, $V_s$, and diluted with other volumes that contain the materials for the test to proceed. We call this final volume the testing volume, $V_t \sim 20\mu l$. These various volumes are then related by:

$$V_t = DV_s = D\sum_{i=1}^{m_j} V_{s;i}^{(j)} = m_j D V_s^{(j)}, \tag{47}$$

where $D$ is the dilution factor. Given that $V_{s;i}^{(j)} \ll V_{o;i}$, the number of RNA molecules in $V_{s;i}^{(j)}$ is a Poisson-distributed random variable, $\mathcal{N}_{R;i}^{(j)}$ of mean:

$$E\mathcal{N}_{R;i}^{(j)} = \tilde{c}_{R;i} V_{s;i}^{(j)} = \tilde{c}_{R;i} V_s^{(j)}. \tag{48}$$

The total number of RNA molecules in the $m_j$-sample pool that is tested is then:

$$N_R^{(j)} = \sum_{i=1}^{m_j} \mathcal{N}_{R;i}^{(j)}. \tag{49}$$

$N_R^{(j)}$ is Poisson distributed with mean:

$$EN_R^{(j)} = V_s^{(j)} \sum_{i=1}^{m_j} \tilde{c}_{R;i} = V_s \frac{1}{m_j} \sum_{i=1}^{m_j} \tilde{c}_{R;i}. \tag{50}$$

Thus, the concentration that can be estimated in the case of an $m_j$-sample pool is:

$$c_R^{(j)} \equiv \frac{1}{m_j} \sum_{i=1}^{m_j} \tilde{c}_{R;i}. \tag{51}$$

Let us now consider an $m_j$-sample pool with $m_j > 1$ that tests positive at the $j$-th stage so that, at stage $j+1$, it is sub-divided into $m_j/m_{j+1}$ sub-pools with $m_{j+1}$ samples each. Given Eq. (47), the size of the volumes that are extracted from the original samples at this stage, $V_s^{(j+1)}$, are related to the previously defined volumes by:

$$V_t = DV_s = m_j DV_s^{(j)} = m_{j+1} DV_s^{(j+1)}. \tag{52}$$

The number of RNA molecules contained in the volume, $V_{s;i}^{(j+1)}$, extracted from the $i$-th original sample is again a Poisson-distributed random variable, $\mathcal{N}_{R;i}^{(j+1)}$, with mean:

$$E\mathcal{N}_{R;i}^{(j+1)} = \tilde{c}_{R;i} V_s^{(j+1)}. \tag{53}$$

The number of RNA molecules in each of the $m_{j+1}$-sample sub-pools is then:

$$N_{R;\ell}^{(j+1)} = \sum_{i=1+(\ell-1)m_{j+1}}^{\ell m_{j+1}} \mathcal{N}_{R;i}^{(j+1)}, \qquad 1 \leq \ell \leq \frac{m_j}{m_{j+1}}, \tag{54}$$

and, as in Eqs.(50)–(51), their means satisfy:

$$EN_{R;\ell}^{(j+1)} = V_s c_{R;\ell}^{(j+1)} \equiv V_s \frac{1}{m_{j+1}} \sum_{i=1+(\ell-1)m_{j+1}}^{\ell m_{j+1}} \tilde{c}_{R;i}, \tag{55}$$

where $c_{R;\ell}^{(j+1)}$ is the RNA concentration that is estimated for the $\ell$-th sub-pool at the end of the ($j+1$-stage) ddPCR test. The sum of the RNA molecules over all the sub-pools that come from the same $m_j$-sample pool is:

$$N_R^{(j+1)} = \sum_{\ell=1}^{m_j/m_{j+1}} N_{R;\ell}^{(j+1)} = \sum_{i=1}^{m_j} \mathcal{N}_{R;i}^{(j+1)}, \tag{56}$$

and, given Eqs. (52), (53), (55) and (56), its mean satisfies:

$$EN_R^{(j+1)} = V_s \sum_{\ell=1}^{m_j/m_{j+1}} c_{R;\ell}^{(j+1)} = V_s^{(j+1)} \sum_{i=1}^{m_j} \tilde{c}_{R;i} = V_s \frac{1}{m_{j+1}} \sum_{i=1}^{m_j} \tilde{c}_{R;i}. \tag{57}$$

Eqs. (51) and (57) imply that:

$$\sum_{\ell=1}^{m_j/m_{j+1}} c_{R;\ell}^{(j+1)} = \frac{m_j}{m_{j+1}} c_R^{(j)}, \tag{58}$$

which, as explained in the paper, can be used to check the self-consistency of the results.

##### 3.3 Formulas to estimate probability from fraction of occurrences.

Here we compile some well established results on the estimation of probabilities from fraction of occurrences, including a discussion on the conditions that guarantee the validity of Eq. (40). Let us assume that an observation can fall into one of two categories, + and −, with probabilities, $P$ and $Q = 1 - P$, respectively. Let us assume that, given $M$ independent observations, a fraction $\hat{P}$, of them falls in the + category. Then, if $M$ is large enough, $\hat{P}$ is a point estimator of $P$ such that:

$$P \in \left[\hat{P} - \zeta_{\alpha/2}\sqrt{\frac{\hat{P}\left(1-\hat{P}\right)}{M}}, \hat{P} + \zeta_{\alpha/2}\sqrt{\frac{\hat{P}\left(1-\hat{P}\right)}{M}}\right], \tag{59}$$

with a $100(1-\alpha)$ confidence level (*i.e.*, there is at least a $100(1-\alpha)\%$ chance that $P$ is contained in the interval given by Eq. (59)) where $\zeta_{\alpha/2}$ is the critical value of the normal distribution for the confidence level [8, 9]. In particular, for $\alpha = 0.05$ (95% confidence) it is $\zeta_{\alpha/2} = 1.96$ and for $\alpha = 0.01$ (99% confidence) it is $\zeta_{\alpha/2} = 2.58$. The large sample approximation holds if $MP \geq 5$ and $MQ \geq 5$ [9] and is most accurate if $0.3 \leq P \leq 0.7$ [8]. Furthermore, under this approximation, the fraction $\hat{P}$ is normally distributed with mean, $P$, and standard deviation, $\sigma_P = \sqrt{PQ/M}$. The confidence interval is sometimes enlarged to include a correction for continuity by adding (subtracting) $1/(2M)$ to the upper (lower) limit of the interval in Eq. (59) [9]. Outside this limit, more complicated expressions can be used for the lower, $P_L$, and upper, $P_U$, limits of the confidence interval that are always valid for a binomially distributed variable:

$$P_L = \frac{x}{x + (M - x + 1)F_{2(M-x+1),2x;\alpha}}, \tag{60}$$

$$P_U = \frac{(x+1)F_{2(x+1),2(M-x);\alpha}}{M - x + (x+1)F_{2(x+1),2(M-x);\alpha}}, \tag{61}$$

with $x = \hat{P}M$ and where the $F_{a,b;\alpha}$ is the $100(1-\alpha)$th percentile of the $F$ distribution with two degrees of freedom [9]. If $MP \geq 5$ and $MQ \geq 5$ but $P < 0.3$ or $P > 0.7$, the lower and upper limits can be computed as:

$$P_L = \frac{2M\hat{P} + \zeta_{\alpha/2}^2 - 1 - \zeta_{\alpha/2}\left(\zeta_{\alpha/2}^2 - \left(2 + \frac{1}{M}\right) + 4\hat{P}\left(M\hat{Q} + 1\right)\right)^{1/2}}{2\left(M + \zeta_{\alpha/2}^2\right)}, \tag{62}$$

$$P_U = \frac{2M\hat{P} + \zeta_{\alpha/2}^2 + 1 + \zeta_{\alpha/2}\left(\zeta_{\alpha/2}^2 + \left(2 - \frac{1}{M}\right) + 4\hat{P}\left(M\hat{Q} + 1\right)\right)^{1/2}}{2\left(M + \zeta_{\alpha/2}^2\right)}, \tag{63}$$

with $\zeta_{\alpha/2}$ as defined before and $\hat{Q} = 1 - \hat{P}$.

### 4 Minimum viral load detectability and maximum pool size using ddPCR

The description introduced in the previous subsection allows us to determine the maximum pool size for which the presence of a single infected sample can be detected with a certain probability. Assuming that ddPCR is able to detect even a single RNA particle, then the presence of only one infected sample in an $m_j$-sample pool will be detected if there is at least one viral RNA molecule in the volume of size, $V_s/m_j$, with which the sample of the infected individual contributes to the total sample volume, $V_s$. Let us assume that the only infected sample of the $m_j$-pool is the $i$-th one. Using the notation and assumptions of the previous subsection, the probability that there is at least one viral RNA molecule in the testing volume is:

$$P(N_R^{(j)} \geq 1) = P(\mathcal{N}_{R;i}^{(j)} \geq 1) = 1 - \exp\left(-\tilde{c}_{R;i}\frac{V_s}{m_j}\right). \tag{64}$$

Setting a lower bound to this probability we can determine the minimum detectable viral concentration for a given $m_j$ or, conversely, the maximum pool size that allows the detection of at least a certain viral RNA concentration. Inserting Eq. (29) in Eq. (64) we obtain the expression that we use in the paper.

### 5 Computation of likelihoods for the detection of flawed samples in pools.

The viability of the tested sample and the quality of the amplification procedure are checked by amplifying the genetic material associated to a piece of human RNA that should be present in any good sample. As described in the paper, we are interested in the possibility of identifying the presence of a single flawed sample in an $m$-sample pool on which a ddPCR test is run. The underlying assumption is that the conditions that guarantee that this distinction is possible would also allow the identification of cases with more than one flawed sample. To this end, we assume that the number of RNA molecules of the $i$-th individual in a volume of size, $V_s$, drawn (with no dilution) from the original sample of the same individual is described by a Poisson distribution with mean $\lambda_i$. The distribution is Poisson as well for any sub-volume of $V_s$ but with mean rescaled by the ratio of volume sizes. Thus, when pooling together $m$ samples whose mean numbers of human RNA molecules in $V_s$ are, respectively, $\lambda_1, \lambda_2, \ldots, \lambda_m$, the number of RNA molecules in $V_s$ of the pool is a Poisson distributed random variable of mean, $\sum_i \lambda_i/m$. The human to human variability is accounted for by assuming that the individual means, $\lambda_i$, of the good samples are actually instances of a Normal distributed random variable of mean $\lambda$ and standard deviation, $\sigma$. Flawed samples, in turn, have $\lambda_i = 0$. We then assume that an $m$-sample pool is characterized by a sequence, $\lambda_1, \lambda_2, \ldots, \lambda_m$ of i.i. (Normal) distributed random variables, if all the samples are good. If there is one flawed sample in the pool, then, the corresponding $\lambda_i$ will be zero and the other $m-1$ will be i.i.d. random variables. We notice that, in both cases, the number of RNA molecules is the same in $V_s$ and in the test volume, $V_t = DV_s$, because the dilution does not change the relevant RNA content.

We now consider that we run the ddPCR test on the pool (subdividing the test volume, $V_t$, into $M$ sub-volumes) and that the number of sub-volumes (nano-droplets) containing no human RNA, $X$, is equal to $x_{obs}$. The two situations that we want to distinguish are: (i) all the individual samples were "good", *i.e.* had $\lambda_i \neq 0$; (ii) one of the samples was "flawed", *i.e.* had $\lambda_i = 0$. We work under the assumption that, if a test satisfies the constraints which guarantee that these two situations are distinguishable, then tests with more than one flawed sample will also be distinguishable from "good" tests. For the comparison we first compute the likelihood of observing $X = x_{obs}$ under both hypotheses. We then compare the likelihoods and compute the critical value, $x_c$, that establishes the border between having one or the other hypothesis as the more probable one given the observations. Finally we compute the accuracy of the classification, *i.e.*, the fraction of instances for which the classification is correct. Given the values, $\lambda_1, \lambda_2, \ldots, \lambda_m$, the probability that there is no RNA in a nano-droplet is $p_{n-f} = \exp(-\sum_i \lambda_i/(Mm))$. Under the assumption that $\lambda_1, \lambda_2, \ldots, \lambda_m$ is a sequence of iid random variables with distribution, $Normal(\lambda, \sigma)$, then $p_{n-f} = e^{-\frac{1}{Mm}Z}$ with $Z \sim Normal(m\lambda, \sqrt{m}\sigma)$. If $m-1$ of the $\lambda_i$s are Normal and one of them is zero, then

$p_{n-f} = e^{-\frac{1}{Mm}Z}$ with $Z \sim Normal((m-1)\lambda, \sqrt{m-1}\sigma)$. Therefore, the likelihoods of obtaining $X = x_{out}$ in each of these situations are:

$$
\begin{aligned}
P(X = x_{obs}|\text{all good}) &= \int_{-\infty}^{\infty} \binom{M}{x_{obs}} e^{-\frac{z}{Mm}x_{obs}} (1 - e^{-\frac{z}{Mm}})^{M-x_{obs}} \phi_{m\lambda,\sqrt{m}\sigma}(z)dz \\
&\approx \binom{M}{x_{obs}} e^{-\lambda x_{obs}/M} (1 - e^{-\lambda/M})^{M-x_{obs}}, \qquad (65) \\
P(X = x_{obs}|\text{one flawed}) &= \int_{-\infty}^{\infty} \binom{M}{x_{obs}} e^{-\frac{z}{Mm}x_{obs}} (1 - e^{-\frac{z}{Mm}})^{M-x_{obs}} \phi_{(m-1)\lambda,\sqrt{m-1}\sigma}(z)dz \\
&\approx \binom{M}{x_{obs}} e^{-\lambda(1-1/m)x_{obs}/M} (1 - e^{-\lambda(1-1/m)/M})^{M-x_{obs}}, \qquad (66)
\end{aligned}
$$

where $\phi_{\mu,\sigma}(z) = \frac{1}{\sqrt{2\pi}\sigma} e^{\frac{(x-\mu)^2}{2\sigma^2}}$ and $\binom{M}{x_{obs}}$ is the corresponding binomial coefficient. Given that the two likelihoods are approximately equal to Binomial distributions centered at different values ($Me^{-\lambda/M}$ and $Me^{-\lambda(1-1/m)/M}$ ), the threshold, $x_c$, can be derived by equating the r.h.s. of Eqs. 65 and 66. We obtain:

$$
x_c = M \left( 1 - \frac{1}{1 - \frac{mM}{\lambda} \left( \log\left(1 - e^{-\lambda(1-1/m)/M}\right) - \log\left(1 - e^{-\lambda/M}\right) \right)} \right). \qquad (67)
$$

If the Binomial distributions can be approximated by Normal ones [2], then a high accuracy is guaranteed if $P(X \leq x_c|\text{one flawed}) \approx P(X \geq x_c|\text{all good})$. Using the normal approximation, the accuracy can then be written as:

$$
\text{Accuracy} = \Phi \left( \frac{x_c - Me^{-\lambda/M}}{\sqrt{M(1 - e^{-\lambda/M})e^{-\lambda/M}}} \right), \qquad (68)
$$

where $\Phi(z) = \int_{\infty}^{z} \phi_{0,1}(x)dx$ with $\phi_{\mu,\sigma}(z)$ as defined before. In the paper we show plots of the accuracy for different pool sizes.

### References


1. Armendáriz I, Ferrari PA, Fraiman D, Martínez JM, Dawson SP. Group testing with nested pools; 2020.

2. Burrows P. Improved estimation of pathogen transmission rates by group testing. Phytopathology. 1987;77(5):363–365.

3. Dube S, Qin J, Ramakrishnan R. Mathematical Analysis of Copy Number Variation in a DNA Sample Using Digital PCR on a Nanofluidic Device. PLOS ONE. 2008;3(8):1–9. doi:10.1371/journal.pone.0002876.

4. Pan Y, Zhang D, Yang P, Poon LLM, Wang Q. Viral load of SARS-CoV-2 in clinical samples. The Lancet Infectious Diseases. 2020;20(4):411 – 412. doi:https://doi.org/10.1016/S1473-3099(20)30113-4.

5. Zheng S, Fan J, Yu F, Feng B, Lou B, Zou Q, et al. Viral load dynamics and disease severity in patients infected with SARS-CoV-2 in Zhejiang province, China, January-March 2020: retrospective cohort study. BMJ. 2020;369. doi:10.1136/bmj.m1443.

6. Liu X, Feng J, Zhang Q, Guo D, Zhang L, Suo T, et al. Analytical comparisons of SARS-COV-2 detection by qRT-PCR and ddPCR with multiple primer/probe sets. Emerging Microbes & Infections. 2020;9(1):1175–1179. doi:10.1080/22221751.2020.1772679.

7. Suo T, Liu X, Feng J, Guo M, Hu W, Guo D, et al. ddPCR: a more accurate tool for SARS-CoV-2 detection in low viral load specimens. Emerging Microbes & Infections. 2020;9(1):1259–1268. doi:10.1080/22221751.2020.1772678.

8. Sheskin DJ. Handbook of Parametric and Nonparametric Statistical Procedures. 5th ed. Chapman & Hall/CRC; 2011.

9. Fleiss JL, Levin B, Paik MC. Statistical Methods for Rates and Proportions. 3rd ed. New York, NY John Wiley & Sons; 2003.
